# PowerAI-CVD – the first Chinese-specific, validated artificial intelligence-powered *in-silico* predictive model for cardiovascular disease

**DOI:** 10.1101/2023.10.08.23296722

**Authors:** Lifang Li, Oscar Hou In Chou, Lei Lu, Hugo Hok Him Pui, Quinncy Lee, Narinder Kaur, Wing Tak Wong, Carlin Chang, Haipeng Liu, Abraham Ka Chung Wai, Bernard Man Yung Cheung, Tong Liu, Gary Tse, Jiandong Zhou

## Abstract

**Background:** The main risk stratification tools for identifying high-risk individuals of cardiovascular disease (CVD) are based on Western populations. Few models are developed specifically for Asian populations and are not enhanced by artificial intelligence (AI). The aim of this study is to develop the first AI-powered quantitative predictive tool for CVD (PowerAI-CVD) incorporate physiological blood pressure measurements, existing diseases and medications, and laboratory tests from Chinese patients.

**Methods:** The study analysed patients who attended family medicine clinics between 1^st^ January 2000 and 31^st^ December 2003. The primary outcome was major adverse cardiovascular events (MACE) defined as a composite of myocardial infarction, heart failure, transient ischaemic attack (TIA)/stroke or cardiovascular mortality, with follow-up until 31^st^ December 2019. The performance of AI-driven models (CatBoost, XGBoost, Gradient Boosting, Multilayer Perceptron, Random Forest, Naïve Bayes, Decision Tree, k-Nearest Neighbor, AdaBoost, SVM-Sigmod) for predicting MACE was compared. Predicted probability (ranging between 0 and 1) of the best model (CatBoost) was used as the baseline *in-silico* marker to predict future MACE events during follow-up.

**Results:** A total of 154,569 patients were included. Over a median follow-up of 16.1 (11.6-17.8) years, 31,061 (20.44%) suffered from MACE (annualised risk: 1.28%). The machine learning *in-silico* marker captured MACE risk from established risk variables (sex, age, mean systolic and diastolic blood pressure, existing cardiovascular diseases, medications (anticoagulants, antiplatelets, antihypertensive drugs, and statins) and laboratory tests (NLR, creatinine, ALP, AST, ALT, HbA1c, fasting glucose, triglyceride, LDL and HDL)). MACE incidences increased quantitatively with ascending quartiles of the *in-silico* marker. The CatBoost model showed the best performance with an area under the receiver operating characteristic curve of 0.869. The CatBoost model based *in-silico* marker shows significant prediction strength for future MACE events, across subgroups (age, sex, prior MACE, etc) and different follow-up durations.

**Conclusions:** The AI-powered risk prediction tool can accurately forecast incident CVD events, allowing personalised risk prediction at the individual level. A dashboard for predictive analytics was developed, allowing real-time dynamic updates of risk estimates from new data. It can be easily incorporated into routine clinical use to aid clinicians and healthcare administrators to identify high-risk patients.

**Graphical Abstract:** 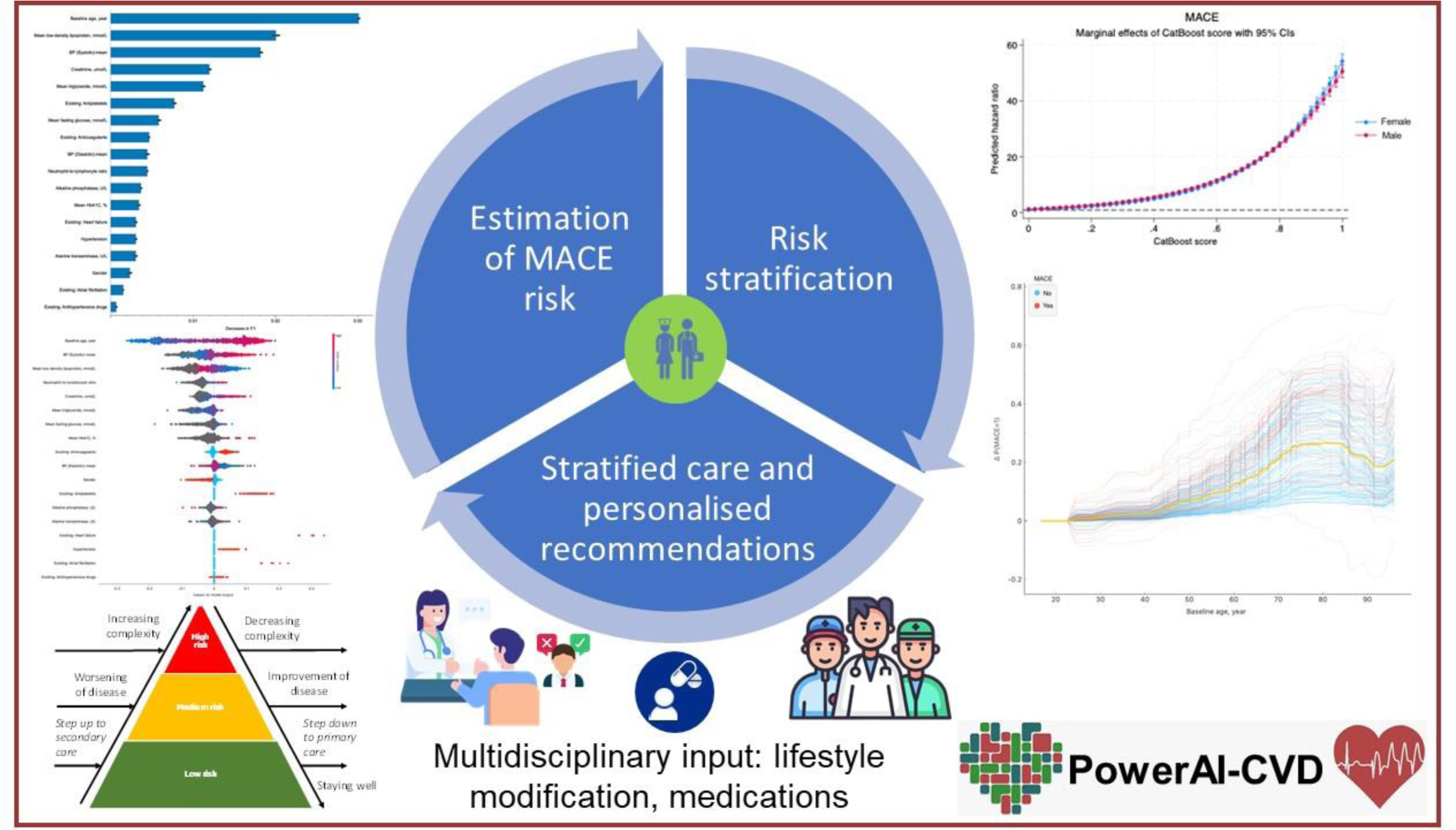

## Introduction

Cardiovascular disease (CVD) is one of the leading causes of deaths globally, leading to health decline and increasing burdens of healthcare costs ^1^. The Global Burden of Diseases, Injuries, and Risk Factors Study (GBD) identified high systolic blood pressure as the leading modifiable risk factor for CVD, accounting for 10.8 million and 11.3 million deaths in 2021 ^2^. In China alone, using the methods of GBD, stroke and ischaemic heart disease were identified as the major causes of death and disability-adjusted life-years (DALYs) ^3^. Therefore, there is a pressing need for effective risk stratification tools that enable clinicians to identify high-risk individuals and recommend treatment accordingly. Of the different tools, the Framingham CVD risk model ^4^, Systematic COronary Risk Evaluation (SCORE) ^5^, pooled cohort equations (PCEs)^6^, and QRisk ^7^were developed using data from Western populations.

However, Asian populations differ in terms of genetic background, environmental exposures and lifestyle choices, disease demographics and epidemiology. For example, Asians have a higher burden of stroke ^8^and hypertension ^9^ but lower burden of hyperlipidaemia ^10^. Whilst the risk factors for CVD are common to both Western and Asian populations, direct application of the above tools can lead to errors in risk estimates ^11^. Moreover, whilst calibration of existing tools is possible ^12–14^, even recalibration of the Framingham CVD risk model led to systematically overestimated CVD risk in older adults from Hong Kong, China ^15^. Yet, few models are specifically developed for Asian populations ^16^. These are Prediction for atherosclerotic CVD Risk in China (China-PAR) ^17^ and absolute risk score from the Japan Arteriosclerosis Longitudinal Study (JALS) ^18^. The limitation is that the China-PAR did not consider non-high-density lipoprotein cholesterol (non-HDL-C) or total cholesterol (TC), which are also important predictors ^19^. Furthermore, at the time of development, the application of artificial intelligence had not yet been popularised ^20^. AI approaches have indeed led to significant improvement in accuracy, sensitivity and specificity compared to regression-based models ^21–23^. Our team was the first to develop AI-based models for predicting adverse outcomes in patients with existing myocardial infarction ^24^, valvular heart disease ^25^, heart failure ^26^ and diabetes ^27,28^. In this study, we develop an AI-driven predictive model for forecasting first and recurrent cardiovascular events using a family medicine clinic cohort from Hong Kong. The novelty is that this model is the first Chinese-specific, validated AI-enhanced model that incorporates physiological blood pressure measurements, existing diseases and medications, and laboratory tests.

## Methods

This study was approved by the Institutional Review Board of the University of Hong Kong/Hospital Authority Hong Kong West Cluster Institutional Review Board (HKU/HA HKWC IRB) (UW-20-250 and UW 23-339) and The Joint Chinese University of Hong Kong (CUHK) Hospital Authority New Territories East Cluster (NTEC) Clinical Research Ethics Committee (CREC) (2018.309 and 2018.643) and complied with the Declaration of Helsinki.

### Study population

This was a retrospective population-based study of prospectively collected electronic health records using the Clinical Data Analysis and Reporting System (CDARS) managed by the Hong Kong Hospital Authority (HA). These records include information from public hospitals, their affiliated outpatient clinics, day-care centres and ambulatory care facilities. This system has been used extensively by local research teams ^29–31^. The inclusion criteria were patients who attended family medicine clinics in the Hong Kong Hospital Authority between 1^st^ January 2000 to 31^st^ December 2003. The exclusion criteria were patients who died within 30 days of the index date or those <18 years old.

### Data extraction

The following clinical and laboratory data were extracted during the baseline period (1^st^ January 2000 to 31^st^ December 2003): demographics (sex and age), blood pressure (mean systolic blood pressure [SBP] and diastolic blood pressure [DBP]), prior comorbidities (diabetes, hypertension, chronic obstructive pulmonary disease, ischaemic heart disease, heart failure, myocardial infarction, and stroke/TIA), medication history and mortality outcomes, which are linked to the local government death registry. The diseases were identified based on International Classification of Diseases (ICD)-9 codes (**Supplementary Appendix Table 1**).

The following laboratory tests at the baseline period (January 1^st^, 2000 to December 31^st^, 2003) were extracted: neutrophil and lymphocyte count, x10^9/L; creatinine, μmol/L; alkaline phosphatase (ALP), U/L; aspartate transaminase, U/L; serum alanine aminotransferase (ALT), U/L; HbA1c, %; fasting glucose, mmol/L; low-density lipoprotein (LDL), high-density lipoprotein (HDL), triglyceride, mmol/L.

### Outcome and statistical analysis

The primary outcome was MACE, defined as any of the following events: myocardial infarction, heart failure, TIA/stroke and cardiovascular mortality, with follow-up until 31^st^ December 2019. Heart failure was included to capture the full spectrum of CVD development^32^. Continuous variables were presented as median (95% confidence interval [CI] or interquartile range [IQR]) and categorical variables were presented as frequency (%). The Mann-Whitney U test was used to compare continuous variables. The χ2 test with Yates’ correction was used for 2×2 contingency data, and Pearson’s χ2 test was used for contingency data for variables with more than two categories.

All significance tests were two-tailed and considered statistically significant when P-values <0.05. Data analyses were performed using RStudio software (Version: 1.1.456) and Python (Version: 3.6). Experiments were simulated on a 15-inch MacBook Pro with 2.2 GHz Intel Core i7 Processor and 16 GB RAM.

### Model development

AL approaches have the advantages of directly considering the relationships and latent interactions between risk variables and outcomes, without requiring assumptions made in the Cox model. In this study, our objective was the development of an *in-silico* predictive marker utilizing machine learning techniques, amalgamating baseline clinical data points of family medicine patients. Our primary aim was to forecast the occurrence of Major Adverse Cardiovascular Events (MACE) during the follow-up period. The predictive model we constructed encompassed a diverse ensemble of machine learning algorithms, including CatBoost ^33^, XGBoost ^34^, Gradient Boosting ^35^, Multilayer Perceptron ^36^, Random Forest ^37^, Naïve Bayes ^38^, Decision Tree ^39^, k-Nearest Neighbor ^40^, AdaBoost ^41^, and SVM-Sigmod model^42^. We also incorporated logistic regression and a dummy classifier as baseline predictors to facilitate comparative analysis. The probability scores from the machine learning model were obtained for each patient as their *in-silico* marker of MACE event, which ranges from 0 (lowest probability) to 1 (highest probability), and was subsequently evaluated as a quantitative marker for future MACE events across subgroups (age, sex, prior MACE, etc) and different follow-up durations. Feature importance was calculated by both AUC reduction approach and SHAP tool ^43^, for cross-checking purpose.

Our workflow involved a random allocation of 80% of cases and an equal number of controls for training purposes, while the remaining 20% of cases and controls were reserved for validation. To mitigate potential sampling bias, this process was reiterated 100 times. To enhance model interpretability and reduce complexity ^44^, we applied the ReliefF feature selection method on the training dataset ^45^, and the resulting feature set was subsequently employed for validation. Additionally, we adopted a 10-fold cross-validation technique, utilizing a grid-search approach for hyperparameter optimization. The resultant predictive model demonstrated its efficacy in identifying MACE status within the validation dataset. Performance metrics were meticulously computed and reported as the mean, accompanied by a 95% confidence interval, across the 100 iterations of our analysis.

## Results

### Basic characteristics of the study cohort

A total of 155,066 patients who attended family medicine clinics managed by the Hong Kong Hospital Authority between 1^st^ January 2000 and 31^st^ December 2003 were included. Of these, 92 patients who died within 30 days after admission and 405 patients under 18 years old at admission were excluded (**Figure 1**). After exclusion, this cohort compromised a total of 154,569 patients (57.54% females, age at diagnosis: 65.85 [Interquartile range: 52.64-76.0 years old]; median follow-up duration: 15.66 years). The baseline characteristics of the cohort are presented in **Table 1**. Amongst the patients, 31,601 developed MACE, 6,704 developed new onset myocardial infarction, 13,826 developed new onset heart failure, and 10,446 developed stroke/TIA. Furthermore, 60,694 patients died during follow-up. The incidence of MACE and all-cause mortality are illustrated in **Supplementary** Figure 1.

**Figure 1.**
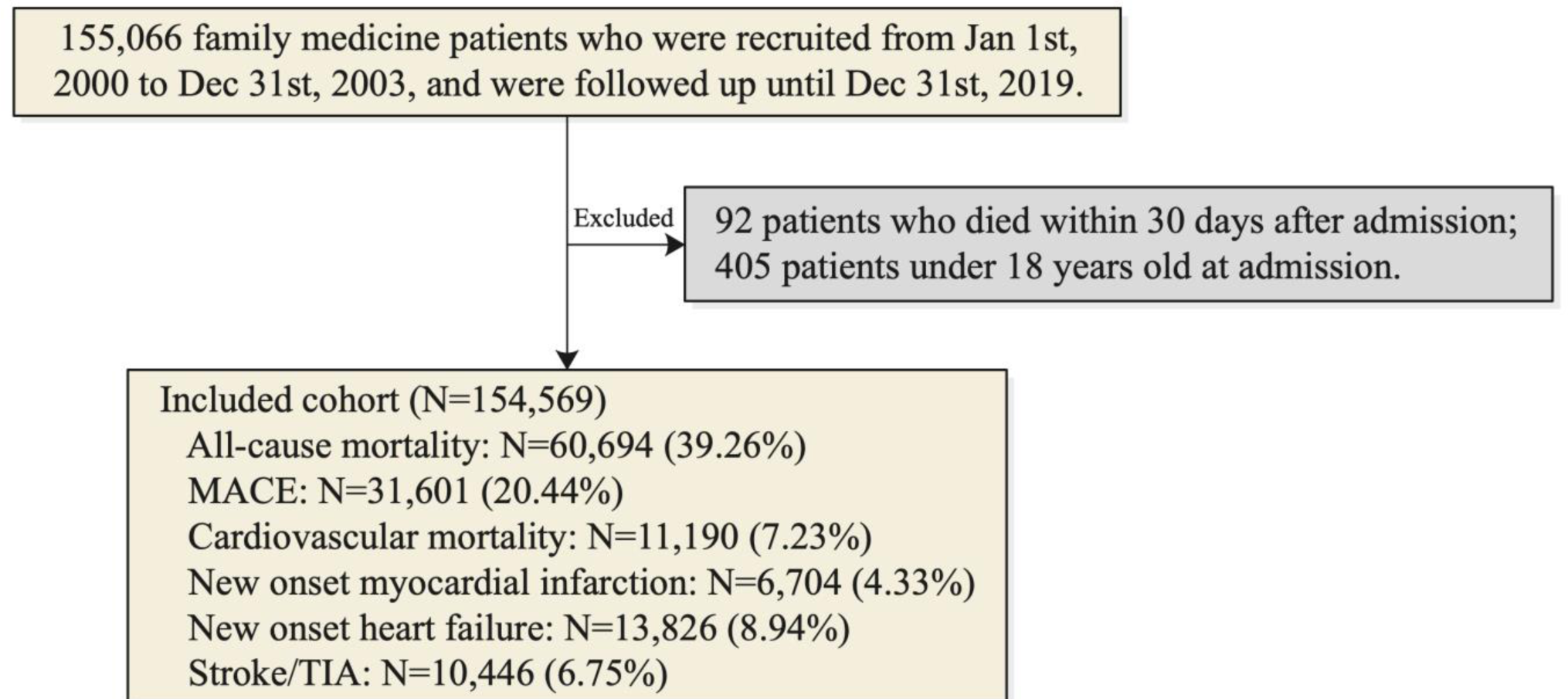
Procedures of data processing. MACE: Major adverse cardiovascular events; TIA: transient ischemic attack.

**Table 1.** Baseline and clinical characteristics of family medicine patients with/without MACE after admission. MACE: Major adverse cardiovascular events; TIA: transient ischemic attack; BP: Blood pressure; IQR: Interquartile range.

| Characteristics | All (N=154569)<br>Median (IQR);N<br>or Count(%) | MACE<br>(N=31601)<br>Median (IQR);N<br>or Count(%) | No MACE<br>(N=122968)<br>Median (IQR);N<br>or Count(%) | P value |
| --- | --- | --- | --- | --- |
| **Outcomes** |  |  |  |  |
| Cardiovascular mortality | 11190(7.23%) | 11190(35.41%) | - | - |
| New onset myocardial infarction | 6704(4.33%) | 6704(21.21%) | - | - |
| New onset heart failure | 13826(8.94%) | 13826(43.75%) | - | - |
| Stroke/TIA | 10446(6.75%) | 10446(33.05%) | - | - |
| **Demographics** |  |  |  |  |
| Male gender | 65627(42.45%) | 13615(43.08%) | 52012(42.29%) | 0.1105 |
| Female gender | 88942(57.54%) | 17986(56.91%) | 70956(57.70%) | 0.1912 |
| Baseline age, year | 65.85(52.64-76.0);n=154569 | 72.08(64.53-78.36);n=31601 | 63.19(50.62-74.86);n=122968 | <0.0001*** |
| **Comorbidities** |  |  |  |  |
| Diabetes mellitus | 10982(7.10%) | 4050(12.81%) | 6932(5.63%) | <0.0001*** |
| Hypertension | 49920(32.29%) | 16313(51.62%) | 33607(27.32%) | <0.0001*** |
| Chronic obstructive pulmonary disease | 721(0.46%) | 248(0.78%) | 473(0.38%) | <0.0001*** |
| Ischemic heart disease | 2245(1.45%) | 1171(3.70%) | 1074(0.87%) | <0.0001*** |
| Heart failure | 1056(0.68%) | 846(2.67%) | 210(0.17%) | <0.0001*** |
| Myocardial infarction | 244(0.15%) | 132(0.41%) | 112(0.09%) | <0.0001*** |
| Atrial fibrillation | 945(0.61%) | 654(2.06%) | 291(0.23%) | <0.0001*** |
| Stroke/TIA | 771(0.49%) | 459(1.45%) | 312(0.25%) | <0.0001*** |
| **Medications** |  |  |  |  |
| Anticoagulants | 64162(41.51%) | 18607(58.88%) | 45555(37.04%) | <0.0001*** |
| Antiplatelets | 14183(9.17%) | 6243(19.75%) | 7940(6.45%) | <0.0001*** |
| Antihypertensive drugs | 33032(21.37%) | 10703(33.86%) | 22329(18.15%) | <0.0001*** |
| Statins | 11043(7.14%) | 3821(12.09%) | 7222(5.87%) | <0.0001*** |
| **Laboratory tests** |  |  |  |  |
| Neutrophil-to-lymphocyte ratio | 2.53(1.74-4.44);n=18392 | 2.92(1.95-5.32);n=5937 | 2.37(1.67-4.06);n=12455 | <0.0001*** |
| Creatinine, umol/L | 84.0(72.0-100.0);n=64104 | 90.0(77.0-107.0);n=18696 | 82.0(71.0-97.0);n=45408 | <0.0001*** |
| Alkaline phosphatase, U/L | 79.0(64.0-96.0);n=35903 | 82.0(67.0-100.0);n=10456 | 77.0(63.0-95.0);n=25447 | <0.0001*** |
| Aspartate transaminase, U/L | 20.0(15.0-29.0);n=25894 | 20.0(14.0-28.0);n=8239 | 21.0(15.0-29.0);n=17655 | <0.0001*** |
| Alanine transaminase, U/L | 20.0(14.0-29.0);n=31129 | 19.0(14.0-28.0);n=9130 | 20.0(14.0-30.0);n=21999 | <0.0001*** |
| BP (Systolic)-mean | 135.41(126.93-143.84);n=111302 | 139.0(131.33-147.29);n=27517 | 134.21(125.5-142.5);n=83785 | <0.0001*** |
| BP (Diastolic)-mean | 73.0(67.5-78.6);n=111302 | 71.74(66.55-77.25);n=27517 | 73.38(67.85-79.0);n=83785 | <0.0001*** |
| Mean HbA1C, % | 7.1(6.33-7.95);n=15483 | 7.2(6.4-8.1);n=5560 | 7.01(6.3-7.9);n=9923 | <0.0001*** |
| Mean fasting glucose, mmol/L | 5.97(5.2-7.57);n=36406 | 6.24(5.35-7.91);n=11848 | 5.8(5.13-7.38);n=24558 | <0.0001*** |
| Mean triglyceride, mmol/L | 1.3(0.97-1.77);n=105550 | 1.29(0.97-1.74);n=26127 | 1.3(0.97-1.79);n=79423 | 0.047* |
| Mean low-density lipoprotein, mmol/L | 2.75(2.31-3.28);n=97844 | 2.58(2.17-3.07);n=24949 | 2.82(2.37-3.34);n=72895 | <0.0001*** |
| Mean high-density lipoprotein, mmol/L | 1.3(1.1-1.56);n=90840 | 1.25(1.05-1.5);n=23942 | 1.32(1.12-1.58);n=66898 | <0.0001*** |

### Comparison of different modelling approaches

The parameters that were significant in predicting specific MACE events in the univariate Cox model were identified as the key variables for inclusion (**Supplementary Table 2**). The variables that predict MACE and their optimal cut-off were identified (**Supplementary Table 3**). The relationship between the laboratory test results with MACE is illustrated in **Supplementary** Figure 2.

The performance of the logistic regression model and the different AI-driven models for the MACE and the specific MACE events were compared. CatBoost had the best performances compared to logistic regression and other AI-driven models in terms of area under the curve (AUC) (AUC: 0.868; 95% Confidence interval [CI]: 0.839-0.939]) (**Table 2**). The order in descending order of AUC: XGBoost, Gradient Boosting, Multilayer Perceptron, Random Forest, Logistic Regression, Naïve Bayes, Decision Tree, k-Nearest Neighbor, AdaBoost, SVM-Sigmod. CatBoost also had the best performances across all models in terms of CA (CA: 0.891; 95% Confidence interval [CI]: 0.856-0.925]), F1-score (F1: 0.836; 95% CI: 0.759-0.89), precision (Precision: 0.826; 95% CI: 0.733-0.889) and recall (Recall: 0.855; 95% CI: 0.842-0.882). The ROC curves for the different prediction models for the adverse outcomes are presented in **Figure 2**. Based on the above observations, the CatBoost model was selected as the most consistent modelling approach.

**Table 2.** Model performance comparisons to predict MACE in family medicine cohort. MACE: Major adverse cardiovascular events; AUC: Area under the curve; CI: Confidence interval.

| Model | AUC [95% CI] | CA [95% CI] | Precision [95% CI] | Recall [95% CI] |
| --- | --- | --- | --- | --- |
| CatBoost | 0.867[0.839-0.938] | 0.895[0.855-0.926] | 0.827[0.735-0.889] | 0.856[0.846-0.882] |
| XGBoost | 0.822[0.802-0.879] | 0.809[0.783-0.818] | 0.779[0.712-0.834] | 0.809[0.726-0.817] |
| Gradient Boosting | 0.811[0.731-0.881] | 0.815[0.759-0.834] | 0.784[0.712-0.795] | 0.814[0.794-0.82] |
| Multilayer Perceptron | 0.776[0.678-0.784] | 0.803[0.799-0.842] | 0.771[0.68-0.837] | 0.803[0.798-0.814] |
| Random Forest | 0.746[0.671-0.807] | 0.792[0.755-0.807] | 0.763[0.685-0.824] | 0.792[0.709-0.849] |
| Logistic Regression | 0.749[0.679-0.838] | 0.803[0.801-0.859] | 0.767[0.702-0.799] | 0.803[0.778-0.902] |
| Naive Bayes | 0.728[0.63-0.788] | 0.711[0.707-0.741] | 0.815[0.806-0.872] | 0.611[0.542-0.664] |
| Decision Tree | 0.702[0.656-0.725] | 0.796[0.767-0.887] | 0.759[0.723-0.769] | 0.796[0.707-0.824] |
| k-Nearest Neighbor | 0.675[0.666-0.758] | 0.778[0.755-0.807] | 0.739[0.682-0.794] | 0.778[0.737-0.796] |
| AdaBoost | 0.635[0.58-0.728] | 0.779[0.778-0.78] | 0.749[0.672-0.789] | 0.779[0.777-0.826] |
| SVM-Sigmoid | 0.575[0.552-0.635] | 0.252[0.209-0.329] | 0.687[0.681-0.715] | 0.252[0.243-0.325] |
| Dummy classifier | 0.555[0.499-0.58] | 0.396[0.374-0.814] | 0.433[0.211-0.694] | 0.396[0.257-0.87] |

**Figure 2.**
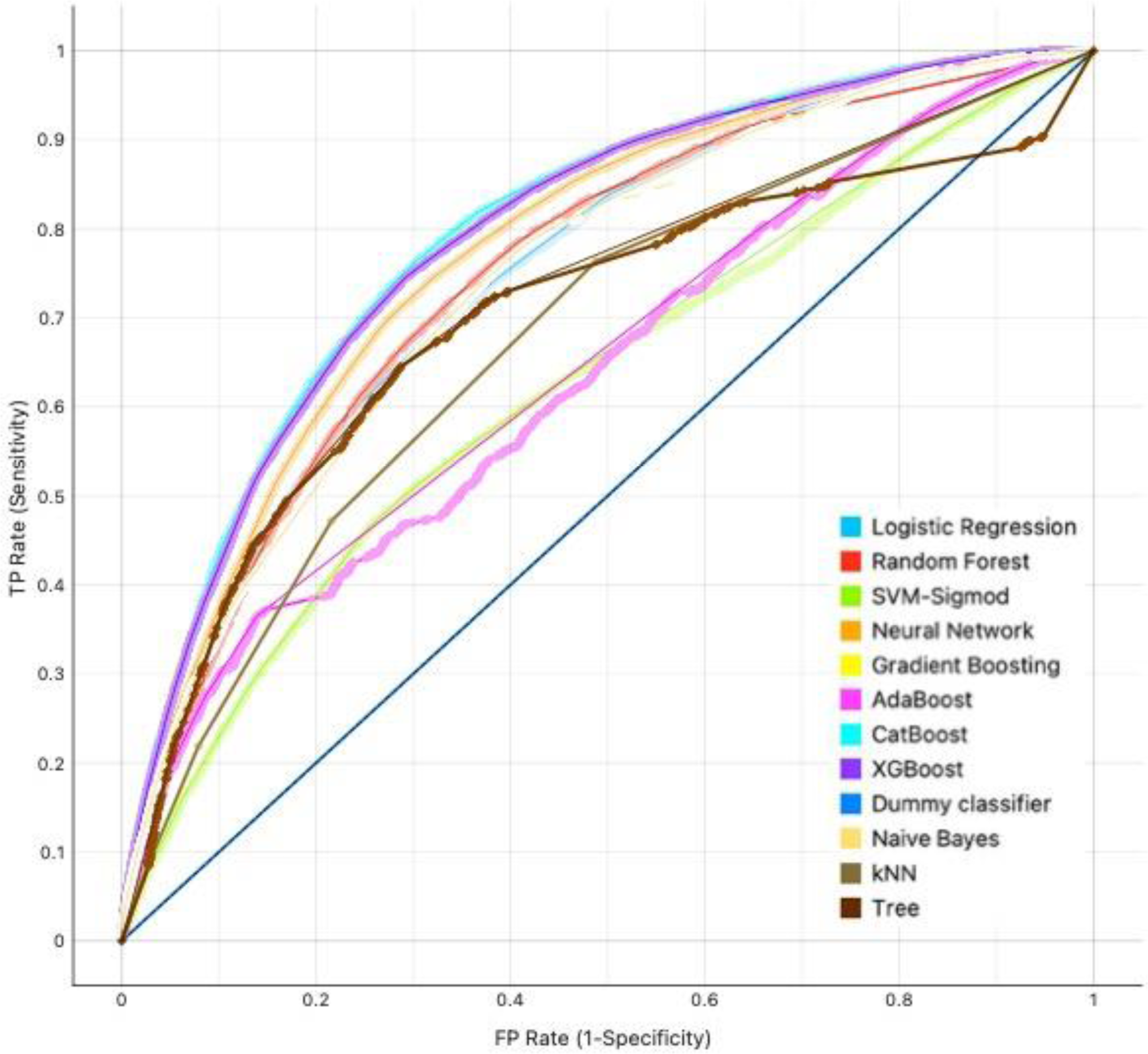
AUROC curve analysis of machine learning algorithms for prediction of MACE in family medicine patients. AUC: Area under the curve; ROC: Receiver operating characteristic; CatBoost: Categorical boosting; XGboost: Extreme gradient boosting; kNN: k-nearest neighbour.

The derived CatBoost *in-silico* marker (**Figure 3a**) demonstrated good clarification ability to predict MACE events in men and women (**Figure 3b**), patients with/without prior MACE (**Figure 3d**), and in subgroups of patients by age admission (**Figure 3e**), stratified by ten deciles. The observed MACE events in each decile of the CatBoost *in-silico* marker was well predicted (**Figure 3c**), with the largest number of MACE events predicted in the 10^th^ decile. The derived CatBoost *in-silico* marker had much better prediction performance for young patients (18-40 years old at admission) and the elderly patients (80+ years old at admission), with average AUC as 0.83 and 0.85 (**Figure 3f**), respectively. And the prediction performance across different age subgroups decreases slightly for those with less than 3-year follow-up duration, while increases afterwards.

**Figure 3.**
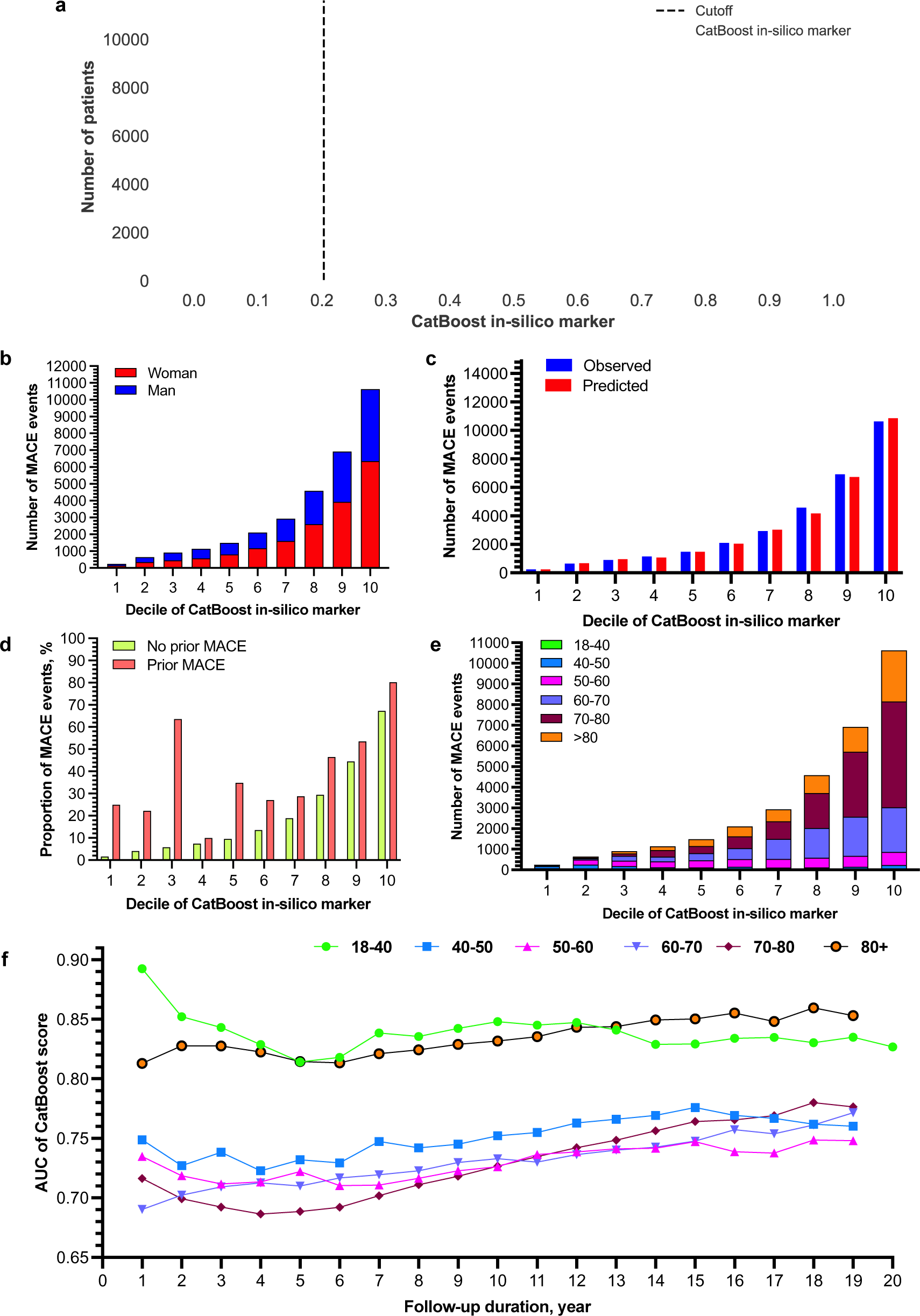
**a**. The number of patients cross the developed CatBoost model based *in-silico* marker and the cutoff. **b**. The number of MACE events amongst patients with different decile of CatBoost risk threshold values stratified by sex. **c**. Observed v.s. predicted number of MACE events by different decile of CatBoost risk threshold values. **d**. The number of MACE events amongst patients with different decile of CatBoost risk threshold values stratified by history of prior MACE. **e**. The number of MACE events amongst patients with different decile of CatBoost risk threshold values stratified by age. **f.** The AUC of the Catboost risk threshold values across the follow-up duration.

### The prediction strength of the CatBoost model based in-silico marker

The distribution analysis demonstrated that the patients with new onset MACE had significantly higher risk threshold values in the CatBoost model compared to patients without new onset MACE (all P values <0.05) (**Supplementary Table 4**). The CatBoost model was evaluated on several partitions of the test population, including sex, age, history of prior MACE, and the follow-up duration. The results demonstrated that the CatBoost model was well-calibrated across both sexes. Meanwhile, the model was more well-calibrated amongst the older age group but less amongst patients younger than 60. Furthermore, the model was more well-calibrated amongst patients without prior MACE events. The predictive strength of the CatBoost model with different follow-up durations was calculated, and it was demonstrated that the risk of MACE prediction was significant across all years (all P value<0.05) (**Supplementary Table 5**). The AUC of the CatBoost model remained consistent across the follow-up durations (**Figure 2e; Supplementary** Figure 3). The predictive strength of the CatBoost model is illustrated in **Figure 4**. The increasing value of derived CatBoost *in-silico* marker indicate much predictive strength of future MACE events (and suboutcomes including cardiovascular mortality, myocardial infarction, heart failure, and stroke/TIA) and all-cause mortality by sex groups, which shows its usefulness in clinical practice as a good risk score system.

**Figure 4.**
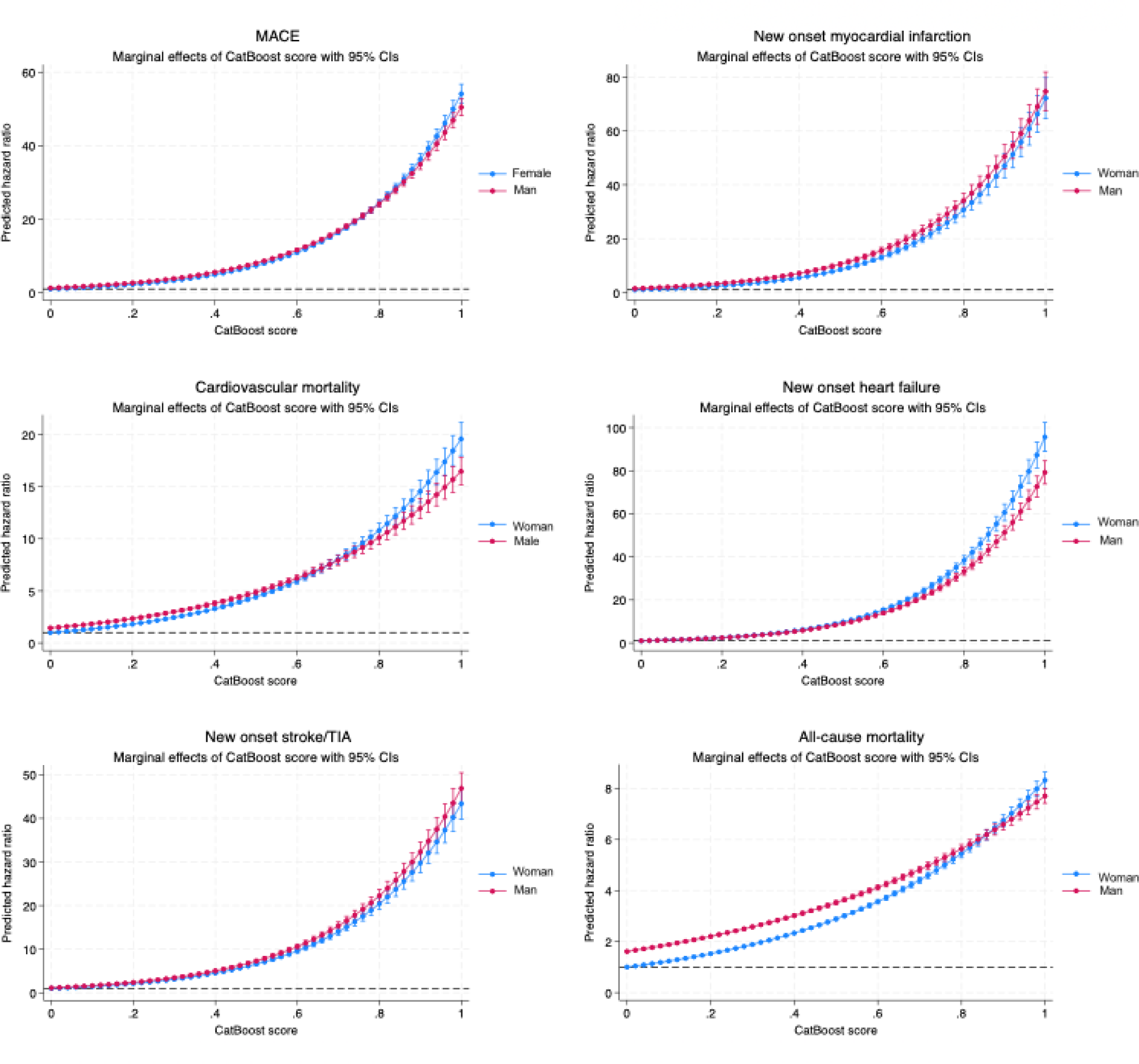
Prediction strength of the developed CatBoost model based *in-silico* marker (predicted probability within [0-1], with baseline and clinical characteristics) to predict future MACE in family medicine patients.

### Important features for MACE using the CatBoost models

The feature importance was calculated using CatBoost to identify the variables with optimal effect when determining the MACE risk. The feature importance indicates how each variable contributes to the model’s accuracy. The 18 most influential predictors are presented in **Figure 5**. According to the result, baseline age was the most important feature of the model. The remaining top five important features included mean low-density lipoprotein, systolic blood pressure, creatinine level and mean triglyceride levels. The SHAP features importance for the CatBoost model was illustrated (**Supplementary** Figure 4). The relationship between age and MACE was demonstrated in the individual conditional expectation plot (**Supplementary figure 5**), which showed that most patients with the higher risk peaked at 80 years old and no obvious interaction was observed.

**Figure 5.**
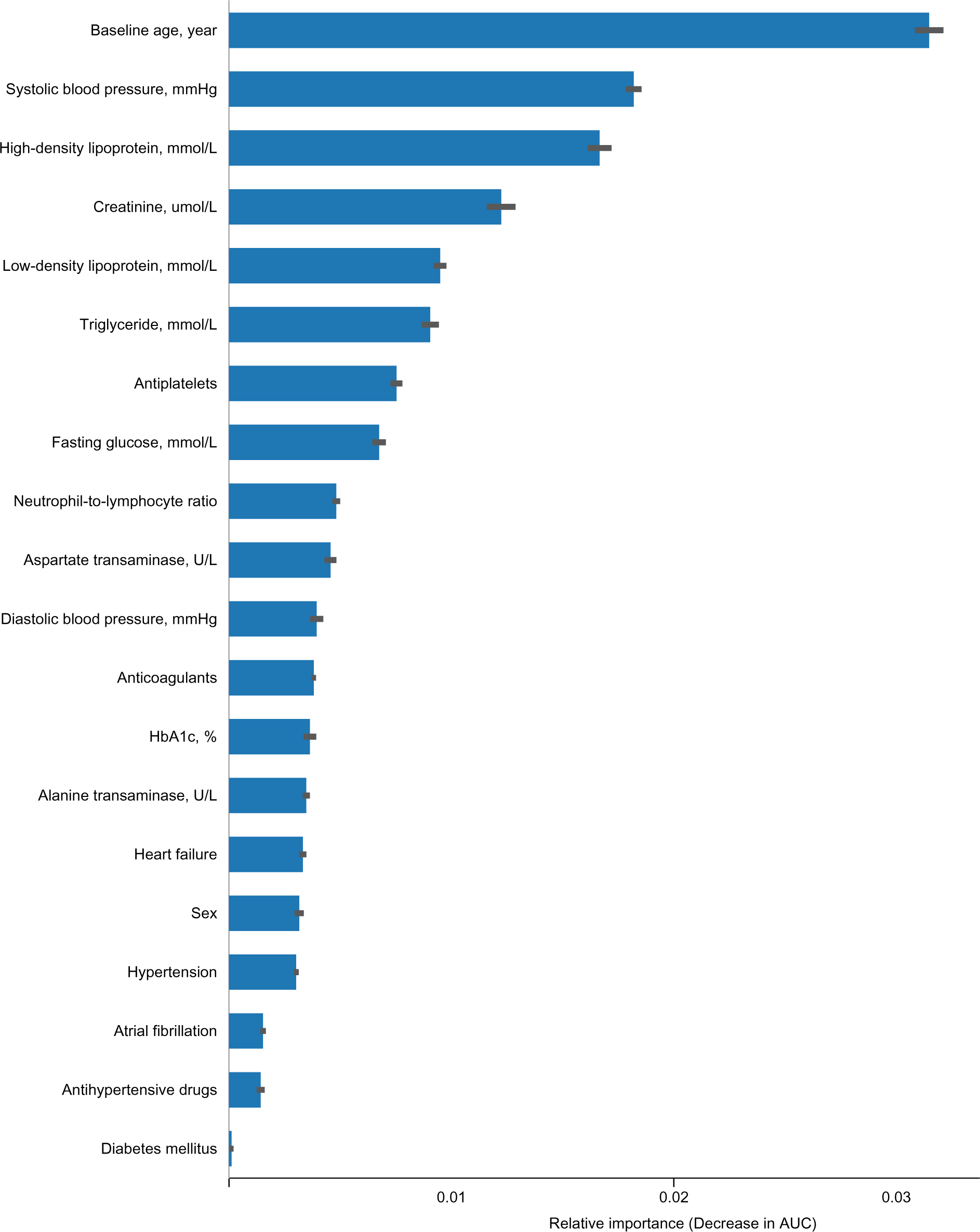
Importance ranking of the different features for the best machine learning model (CatBoost) to predict MACE in Hong Kong Chinese patients attending public family medicine clinics.

### Dashboard for Model implementation

The PowerAI-CVD model supports clinical decision making by presenting key information on a dashboard (**Figure 6**). The most important input variables from current diseases, blood pressure, laboratory tests and existing medications are used for calculating the risk of MACE at 3, 5, 10 and 20 years. The trends of the different risk factors can aid clinicians and patients to monitor the effectiveness of ongoing lifestyle modifications and/or pharmacotherapy. The value of this dashboard is that appropriate recommendations for antihypertensive and lipid lowering drugs are available, with targets of blood pressure, lipid and glucose control. These functions can serve to facilitate guideline-directed treatment and improve adherence to best current practices to improve patient outcomes.

**Figure 6.**
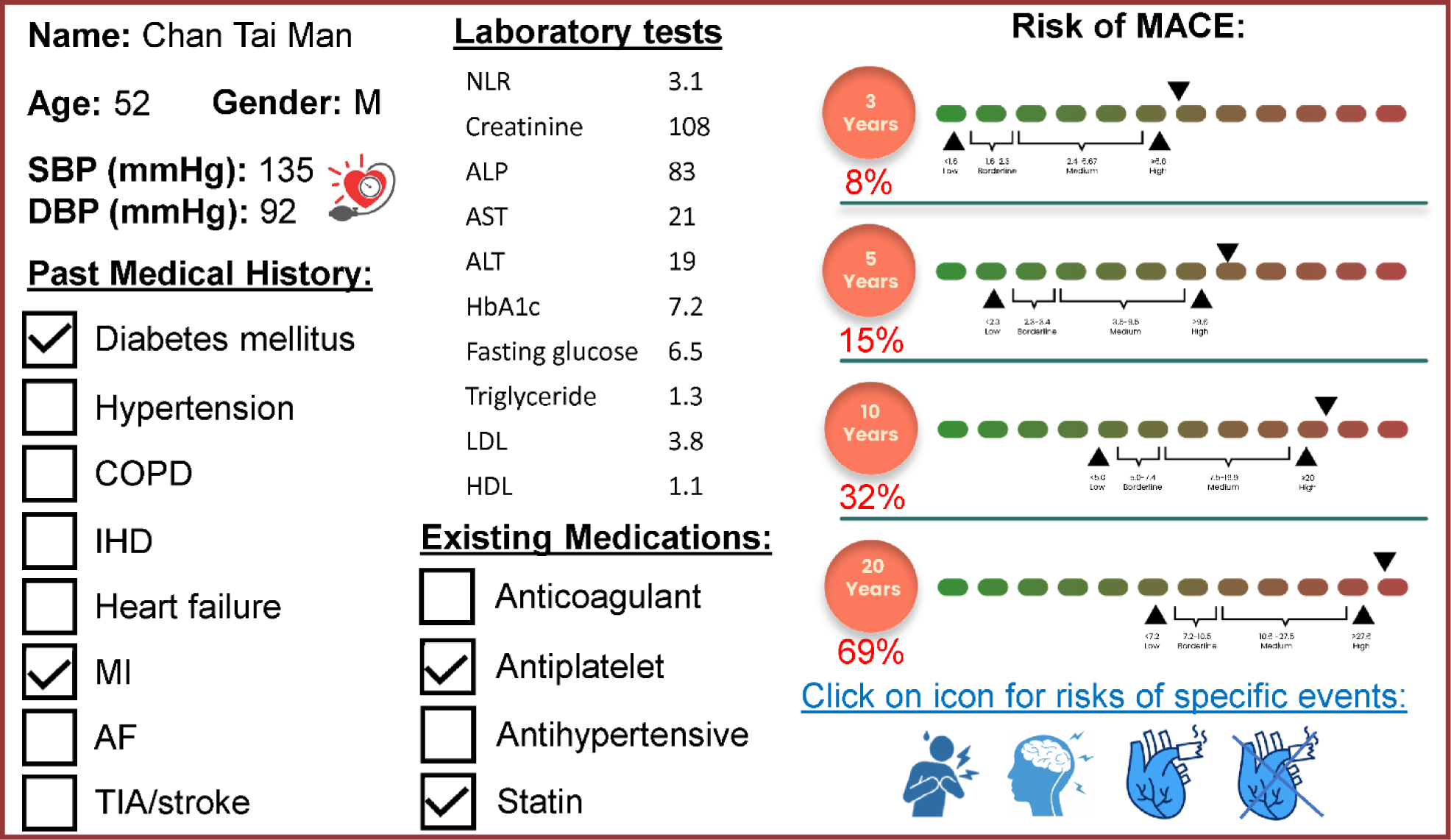
Model implementation. The dashboard shows the real-time risks using physiological blood pressure measurements, disease status, medications and laboratory findings.

## Discussion

In this study, we developed the first AI-powered CVD model (PowerAI-CVD) to incorporate physiological blood pressure measurements, existing diseases and medications, and laboratory tests for predicting incident MACE using a big data approach. The main findings are that CatBoost significantly outperformed other machine learning and regression-based models.

Current risk models for CVD have been developed using Western cohort data. Yet, Chinese subjects have differing risks of different adverse events owing to differences in genetics, environment exposure, and disease status. For example, for stroke and TIA, the intracranial atherosclerosis more prevalent as an aetiology of stroke amongst Chinese^46^. Of the different risk factors, our analysis identified age, LDL and systolic blood pressure as the 3 most important risk factors. These findings are in line with the notion that age is the most determinant of a person’s cardiovascular health ^47^. For LDL, its concentration in the bloodstream and longer duration of exposure are both important predictive factors of CVD risk ^48^. Blood pressure influence local hemodynamics in the brain and the heart, leading to a higher translesional pressure gradient which is directly associated with risk of plaque rupture.

Accurate risk estimates are required to identify high-risk individuals who can benefit from treatment and prevent overtreatment of low-risk individuals. Given that prevention is better than cure, our tool can be used in primary care settings. A limited number of CVD risk models based on Asian cohort data are available ^49^. These have the major limitations of not having undergone validation, and not AI driven. By contrast, our team overcomes both limitations by developing the first AI-powered risk model for predicting new CVD events using territory-wide data from Hong Kong. Such a big data approach has the advantage of being highly generalisable as it is based on the entire population of Hong Kong, with the ability of analysing multiple clinically relevant outcomes. Our model is capable of considering new information with dynamic risk assessment ^50^. For example, with updated blood pressure and laboratory measurements, our dashboard can provide updated risks to the clinicians.

### Clinical implications

The majority of the risk factors are modifiable, and by extension, CVD events are largely preventable. The predictors used in our PowerAI-CVD model do not require specialised testing, but rather demographics, disease status, medication history and routine laboratory parameters reflecting inflammatory, renal, liver, glycemic and lipid levels. The AI-powered risk prediction tool can accurately forecast incident CVD events, allowing personalised risk prediction at the individual level. A dashboard for predictive analytics was developed, allowing real-time dynamic updates of risk estimates from new data. Patients are also empowered as the dashboard provides information that is easy-to-understand. The model can thus be easily incorporated into routine clinical use to aid clinicians and healthcare administrators to identify high-risk patients. Personalised recommendations for lifestyle changes and suggested medications are provided, facilitating decision making and collaborative care between patients and their clinicians. In the long-run, implementation of this tool can reduce healthcare costs at the systems level, enabling sustainable development and healthy aging.

### Limitations

As the aim of this the model was to provide long-term (>10 year) risk estimates, treatment effects from newer pharmacological agents such as proprotein convertase subtilisin/kexin type 9 (PCSK9) inhibitors and sodium-glucose cotransporter-2 (SGLT2) inhibitors are not considered. Future studies are underway to incorporate effects of these medication classes on CVD risk. Moreover, our team has previously reported the incremental value of incorporating visit-to-visit variability in blood pressure, lipid and glycaemic test results for improving risk stratification. Our subsequent studies will incorporate automated methods of considering such measures of variability to calculate not only the estimated future risks, but also likely trajectory of the risk estimates.

## Conclusion

The first ever Chinese-specific, validated, CVD risk model with AI enhancement (PowerAI-CVD) was developed using a family medicine cohort of Chinese patients. It can accurately forecast incident CVD events, allowing personalised risk prediction at the individual level. A dashboard for predictive analytics was developed, allowing real-time dynamic updates of risk estimates from new data. It can be easily incorporated into routine clinical use to aid clinicians and healthcare administrators to identify high-risk patients.

## Ethical approval statement

This study was approved by the Institutional Review Board of the University of Hong Kong/Hospital Authority Hong Kong West Cluster (HKU/HA HKWC IRB) (UW-20-250) and complied with the Declaration of Helsinki.

## Availability of data and materials

An anonymised version without identifiable or personal information is available from the corresponding authors upon reasonable request for research purposes.

## Guarantor Statement

All authors approved the final version of the manuscript. GT is the guarantor of this work and, as such, had full access to all the data in the study and takes responsibility for the integrity of the data and the accuracy of the data analysis.

## Author contributions

Data analysis: Lifang Li, Jiandong Zhou

Data review: Lei Lu, Gary Tse

Data acquisition: Carlin Chang, Teddy Tai Loy Lee, Hugo Hok Him Pui, Bosco Kwok Hei Leung

Data interpretation: All authors

Drafting of manuscript: Lifang Li, Oscar Chou, Gary Tse, Tong Liu, Jiandong Zhou Critical revision of manuscript: All authors

Study supervision: Carlin Chang, Abraham Ka Chung Wai, Bernard Man Yung Cheung, Gary Tse, Jiandong Zhou

## Data Availability

Anonymised dataset produced in the present study are available upon reasonable request to the authors

## Acknowledgements

This analysis uses data or information from the Harmonized CHARLS dataset and Codebook, Version D as of June 2021 developed by the Gateway to Global Aging Data. The development of the Harmonized CHARLS was funded by the National Institute on Aging (R01 AG030153, RC2 AG036619, R03 AG043052). For more information, please refer to https://g2aging.org/. The Authors are also grateful to The Chinese Longitudinal Healthy Longevity Survey (CLHLS) Team for making their datasets available to our team.

## References

1. Vaduganathan M, Mensah George A, Turco Justine V, Fuster V, Roth Gregory A. The Global Burden of Cardiovascular Diseases and Risk. Journal of the American College of Cardiology. 2022;80(25):2361–2371.

2. Razo C, Welgan CA, Johnson CO, et al. Effects of elevated systolic blood pressure on ischemic heart disease: a Burden of Proof study. Nat Med. 2022;28(10):2056–2065.

3. Zhou M, Wang H, Zeng X, et al. Mortality, morbidity, and risk factors in China and its provinces, 1990-2017: a systematic analysis for the Global Burden of Disease Study 2017. Lancet. 2019;394(10204):1145–1158.

4. D’Agostino RB, Sr., Vasan RS, Pencina MJ, et al. General cardiovascular risk profile for use in primary care: the Framingham Heart Study. Circulation. 2008;117(6):743–753.

5. Conroy RM, Pyorala K, Fitzgerald AP, et al. Estimation of ten-year risk of fatal cardiovascular disease in Europe: the SCORE project. Eur Heart J. 2003;24(11):987–1003.

6. Goff DC, Jr., Lloyd-Jones DM, Bennett G, et al. 2013 ACC/AHA guideline on the assessment of cardiovascular risk: a report of the American College of Cardiology/American Heart Association Task Force on Practice Guidelines. Circulation. 2014;129(25 Suppl 2):S49–73.

7. Hippisley-Cox J, Coupland C, Vinogradova Y, Robson J, May M, Brindle P. Derivation and validation of QRISK, a new cardiovascular disease risk score for the United Kingdom: prospective open cohort study. BMJ. 2007;335(7611):136.

8. Irawati S, Wasir R, Floriaan Schmidt A, et al. Long-term incidence and risk factors of cardiovascular events in Asian populations: systematic review and meta-analysis of population-based cohort studies. Curr Med Res Opin. 2019;35(2):291–299.

9. Mills KT, Bundy JD, Kelly TN, et al. Global Disparities of Hypertension Prevalence and Control: A Systematic Analysis of Population-Based Studies From 90 Countries. Circulation. 2016;134(6):441–450.

10. Asia Pacific Cohort Studies C, Barzi F, Patel A, et al. Cardiovascular risk prediction tools for populations in Asia. J Epidemiol Community Health. 2007;61(2):115–121.

11. Liau SY, Mohamed Izham MI, Hassali MA, Shafie AA. A literature review of the cardiovascular risk-assessment tools: applicability among Asian population. Heart Asia. 2010;2(1):15–18.

12. Van Calster B, McLernon DJ, van Smeden M, et al. Calibration: the Achilles heel of predictive analytics. BMC Med. 2019;17(1):230.

13. Lee CH, Woo YC, Lam JK, et al. Validation of the Pooled Cohort equations in a long-term cohort study of Hong Kong Chinese. J Clin Lipidol. 2015;9(5):640–646 e642.

14. Liu J, Hong Y, D’Agostino RB, Sr., et al. Predictive value for the Chinese population of the Framingham CHD risk assessment tool compared with the Chinese Multi-Provincial Cohort Study. JAMA. 2004;291(21):2591–2599.

15. Leung JY, Lin SL, Lee RS, Lam TH, Schooling CM. Framingham risk score for predicting cardiovascular disease in older adults in Hong Kong. Hong Kong Med J. 2018;24 Suppl 4(4):8–11.

16. Zhang Y, Miao H, Chia YC, et al. Cardiovascular risk assessment tools in Asia. J Clin Hypertens (Greenwich). 2022;24(4):369–377.

17. Yang X, Li J, Hu D, et al. Predicting the 10-Year Risks of Atherosclerotic Cardiovascular Disease in Chinese Population: The China-PAR Project (Prediction for ASCVD Risk in China). Circulation. 2016;134(19):1430–1440.

18. Harada A, Ueshima H, Kinoshita Y, et al. Absolute risk score for stroke, myocardial infarction, and all cardiovascular disease: Japan Arteriosclerosis Longitudinal Study. Hypertens Res. 2019;42(4):567–579.

19. Li HH, Huang S, Liu XZ, Zou DJ. Applying the China-PAR Risk Algorithm to Assess 10-year Atherosclerotic Cardiovascular Disease Risk in Populations Receiving Routine Physical Examinations in Eastern China. Biomed Environ Sci. 2019;32(2):87–95.

20. Sundström J, Schön TB. Machine Learning in Risk Prediction. Hypertension. 2020;75(5):1165–1166.

21. Chiarito M, Luceri L, Oliva A, Stefanini G, Condorelli G. Artificial Intelligence and Cardiovascular Risk Prediction: All That Glitters is not Gold. Eur Cardiol. 2022;17:e29.

22. Sun X, Yin Y, Yang Q, Huo T. Artificial intelligence in cardiovascular diseases: diagnostic and therapeutic perspectives. European Journal of Medical Research. 2023;28(1):242.

23. Forrest IS, Petrazzini BO, Duffy A, et al. Machine learning-based marker for coronary artery disease: derivation and validation in two longitudinal cohorts. Lancet. 2023;401(10372):215–225.

24. Li CK, Xu Z, Ho J, et al. Association of NPAC score with survival after acute myocardial infarction. Atherosclerosis. 2020;301:30–36.

25. Tse G, Zhou J, Lee S, et al. Multi-parametric system for risk stratification in mitral regurgitation: A multi-task Gaussian prediction approach. Eur J Clin Invest. 2020;50(11):e13321.

26. Tse G, Zhou J, Woo SWD, et al. Multi-modality machine learning approach for risk stratification in heart failure with left ventricular ejection fraction 45. ESC Heart Fail. 2020.

27. Lee S, Zhou J, Wong WT, et al. Glycemic and lipid variability for predicting complications and mortality in diabetes mellitus using machine learning. BMC Endocr Disord. 2021;21(1):94.

28. Lee S, Zhou J, Leung KSK, et al. Development of a predictive risk model for all-cause mortality in patients with diabetes in Hong Kong. BMJ Open Diabetes Res Care. 2021;9(1).

29. Lee S, Zhou J, Li KHC, et al. Territory-wide cohort study of Brugada syndrome in Hong Kong: predictors of long-term outcomes using random survival forests and non-negative matrix factorisation. Open Heart. 2021;8(1).

30. Liu Y, Ling L, Wong SH, et al. Outcomes of respiratory viral-bacterial co-infection in adult hospitalized patients. EClinicalMedicine. 2021;37:100955.

31. Zhou J, Lee S, Wang X, et al. Development of a multivariable prediction model for severe COVID-19 disease: a population-based study from Hong Kong. NPJ Digit Med. 2021;4(1):66.

32. Tsai IT, Wang C-P, Lu Y-C, et al. The burden of major adverse cardiac events in patients with coronary artery disease. BMC Cardiovascular Disorders. 2017;17(1):1.

33. Ostroumova L, Gusev G, Vorobev A, Dorogush AV, Gulin A. CatBoost: unbiased boosting with categorical features. Paper presented at: Neural Information Processing Systems 2017.

34. Chen T, Guestrin C. XGBoost: A Scalable Tree Boosting System. 2016.

35. Friedman JH. Greedy Function Approximation: A Gradient Boosting Machine. The Annals of Statistics. 2001;29(5):1189–1232.

36. Ramchoun H, Idrissi MAJ, Ghanou Y, Ettaouil M. Multilayer Perceptron: Architecture Optimization and Training. Int J Interact Multim Artif Intell. 2016;4:26–30.

37. Biau G, Scornet E. A random forest guided tour. TEST. 2016;25(2):197–227.

38. Rish I. An Empirical Study of the Naïve Bayes Classifier. IJCAI 2001 Work Empir Methods Artif Intell. 2001;3.

39. Myles AJ, Feudale RN, Liu Y, Woody N, Brown SD. An introduction to decision tree modeling. Journal of Chemometrics. 2004;18.

40. Peterson L. K-nearest neighbor. Scholarpedia. 2009;4:1883.

41. Bernhard S, John P, Thomas H. AdaBoost is Consistent. In: Advances in Neural Information Processing Systems 19: Proceedings of the 2006 Conference. MIT Press; 2007:105–112.

42. Lin H-T, Lin C-J. A Study on Sigmoid Kernels for SVM and the Training of non-PSD Kernels by SMO-type Methods. Neural Computation. 2003.

43. Slack D, Hilgard S, Jia E, Singh S, Lakkaraju H. Fooling LIME and SHAP: Adversarial Attacks on Post hoc Explanation Methods. 2020.

44. Rajkomar A, Dean J, Kohane I. Machine Learning in Medicine. New England Journal of Medicine. 2019;380(14):1347–1358.

45. Kononenko I, Šimec E, Robnik-Šikonja M. Overcoming the Myopia of Inductive Learning Algorithms with RELIEFF. Applied Intelligence. 1997;7(1):39–55.

46. Leng X, Lan L, Ip HL, et al. Hemodynamics and stroke risk in intracranial atherosclerotic disease. Ann Neurol. 2019;85(5):752–764.

47. North BJ, Sinclair DA. The intersection between aging and cardiovascular disease. Circ Res. 2012;110(8):1097–1108.

48. Domanski MJ, Tian X, Wu CO, et al. Time Course of LDL Cholesterol Exposure and Cardiovascular Disease Event Risk. J Am Coll Cardiol. 2020;76(13):1507–1516.

49. Zhiting G, Jiaying T, Haiying H, Yuping Z, Qunfei Y, Jingfen J. Cardiovascular disease risk prediction models in the Chinese population-a systematic review and meta-analysis. BMC Public Health. 2022;22(1):1608.

50. Teramukai S, Okuda Y, Miyazaki S, Kawamori R, Shirayama M, Teramoto T. Dynamic prediction model and risk assessment chart for cardiovascular disease based on on-treatment blood pressure and baseline risk factors. Hypertension Research. 2016;39(2):113–118.

