## Supplementary Appendix for "PowerAI-CVD – the first Chinese-specific, validated artificial intelligence-powered *in-silico* predictive model for cardiovascular disease"

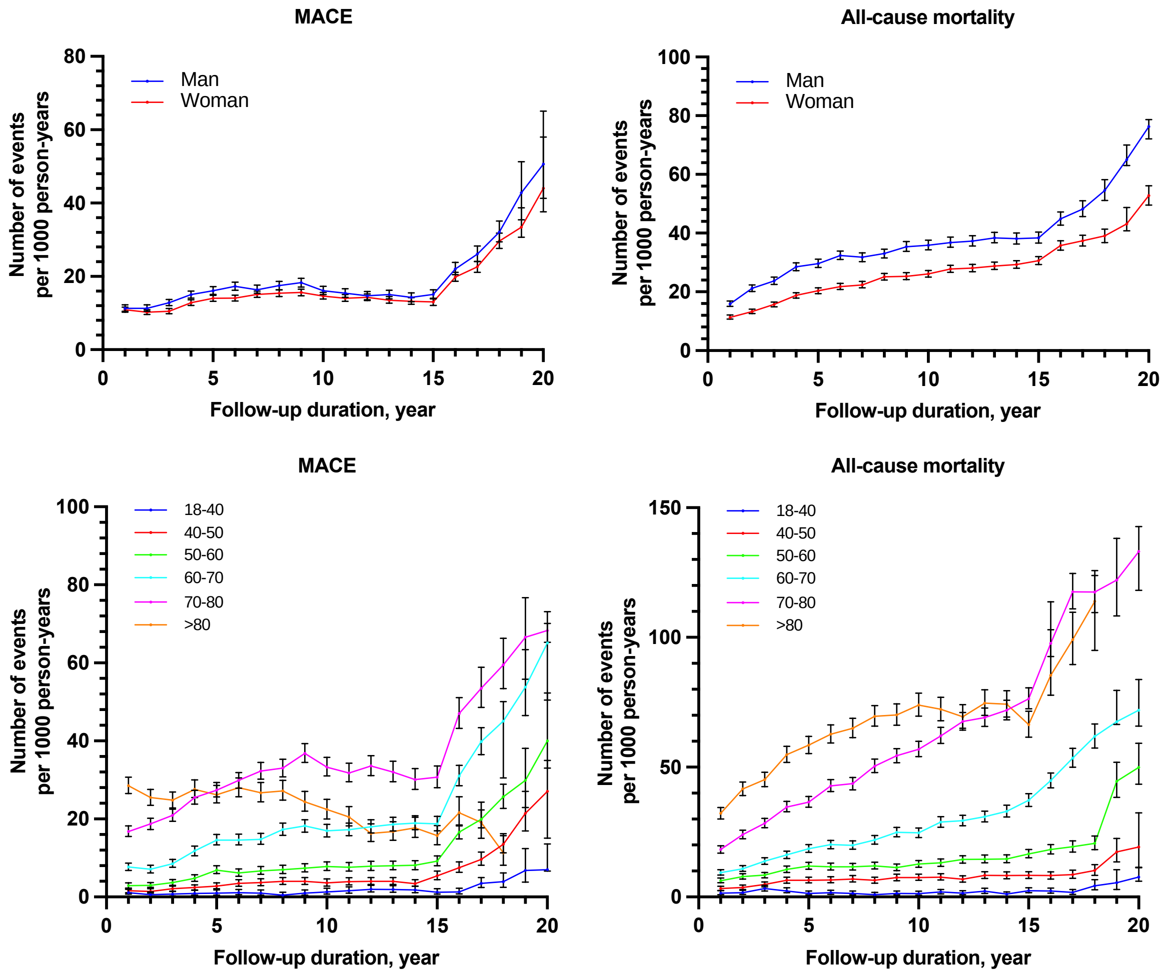

**Supplementary Figure 1.** Incidence of the MACE and all-cause mortality (number of events per 1000 patient-years) stratified by gender and age at baseline.

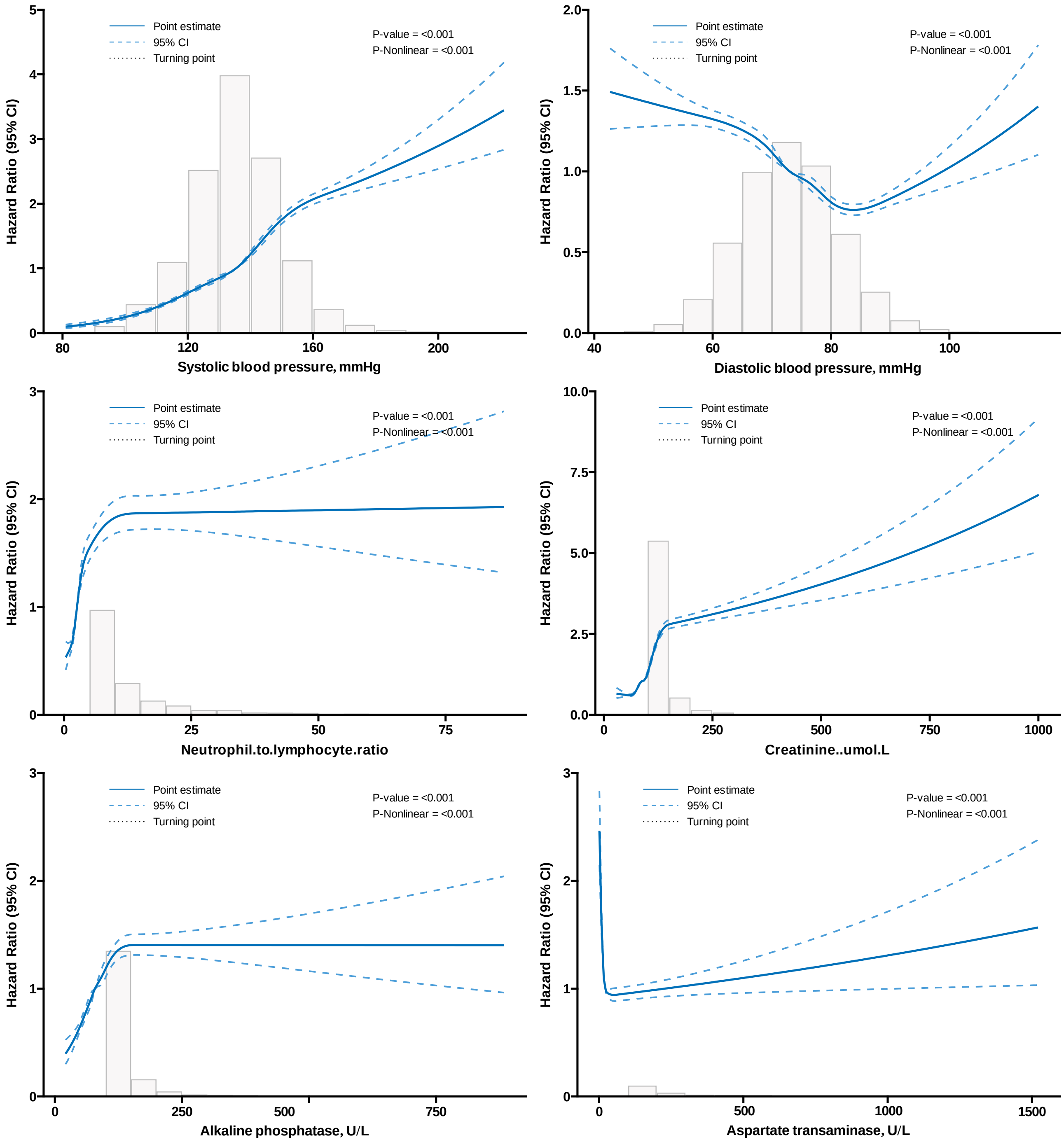

**Supplementary Figure 2a.** Investigating the relationship between laboratory tests and MACE in patients attending family medicine clinics, using restricted cubic splines approach. Model with 7 knots located at 2.5th, 18.33rd, 34.17th, 50th, 65.83th, 81.67th, and 97.5th percentiles.

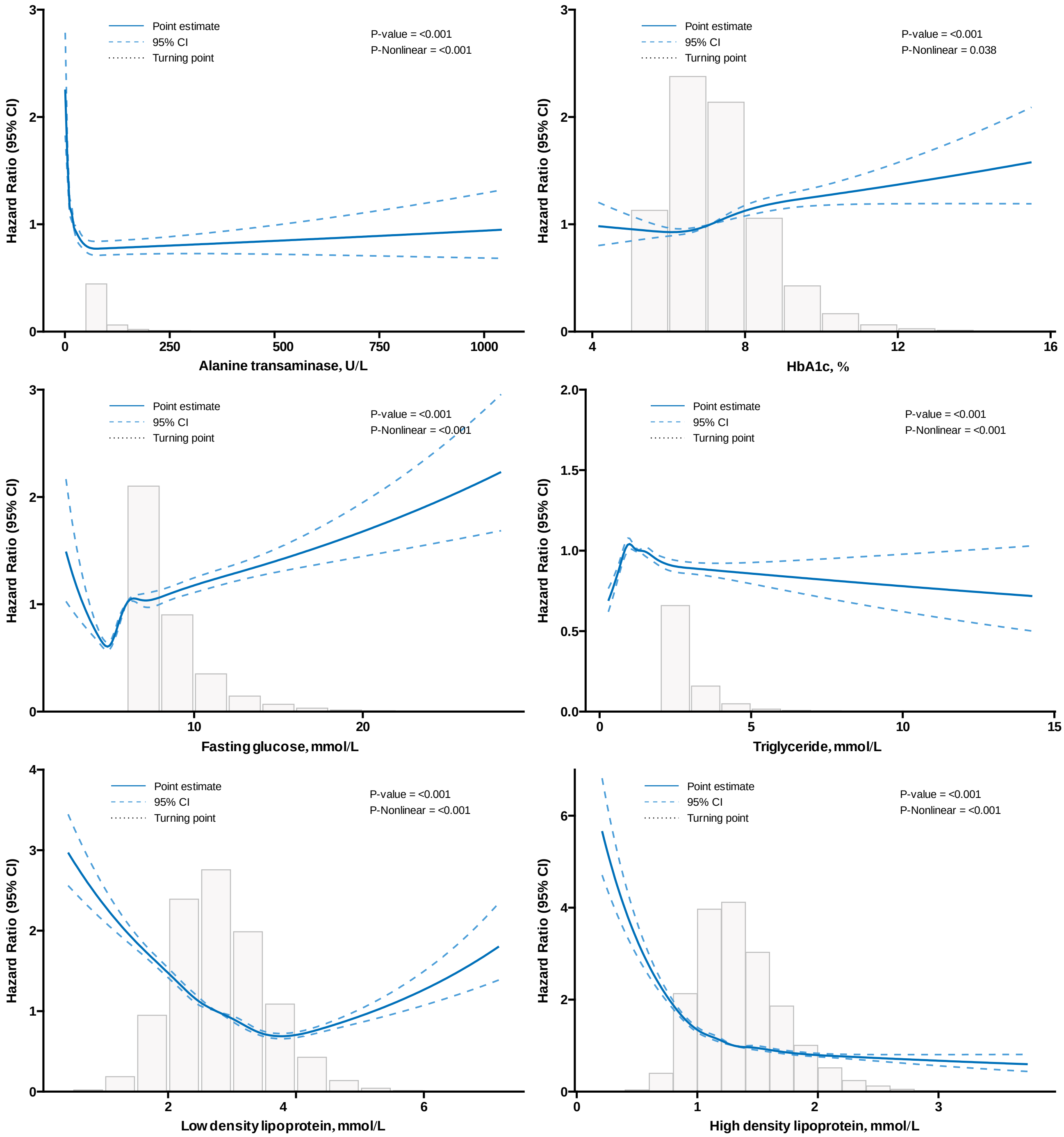

**Supplementary Figure 2b.** Investigating the relationship between laboratory tests and MACE in patients attending family medicine clinics, using restricted cubic splines function approach.

Model with 7 knots located at 2.5th, 18.33th, 34.17th, 50th, 65.83th, 81.67th, and 97.5th percentiles.

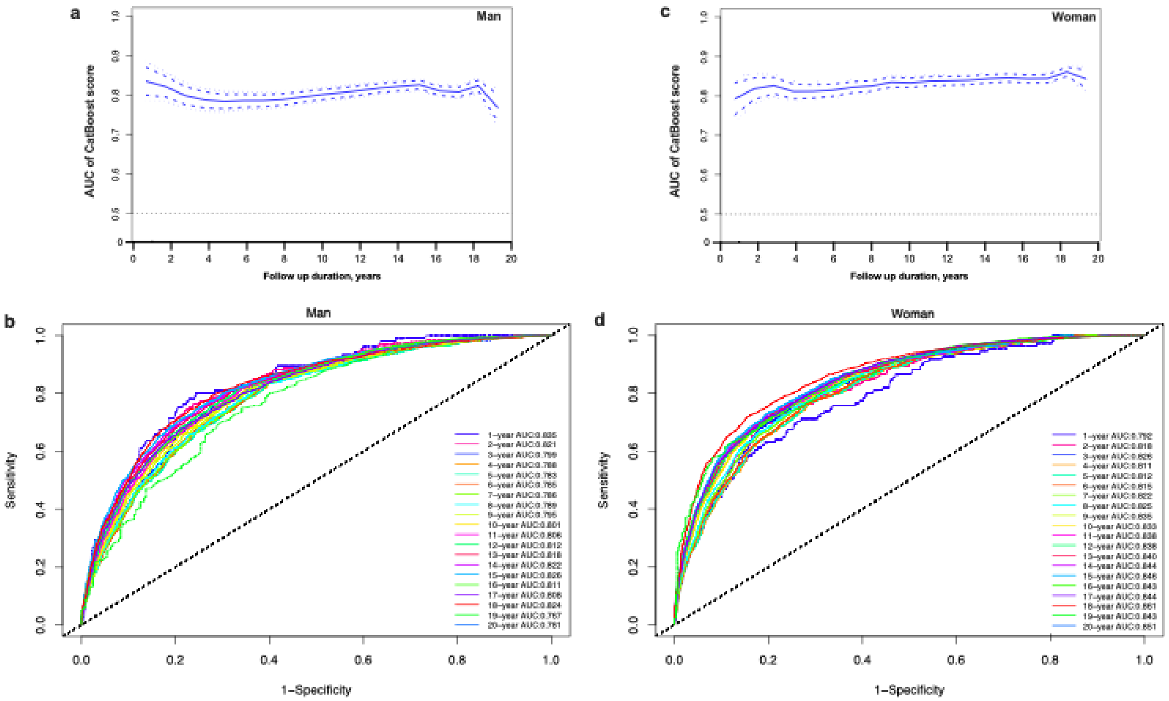

**Supplementary Figure 3.** a, c. The relationship between the AUC of CatBoost score and the follow-up duration stratified by gender; b, d. the AUC curve stratified by the follow-up duration and the gender.

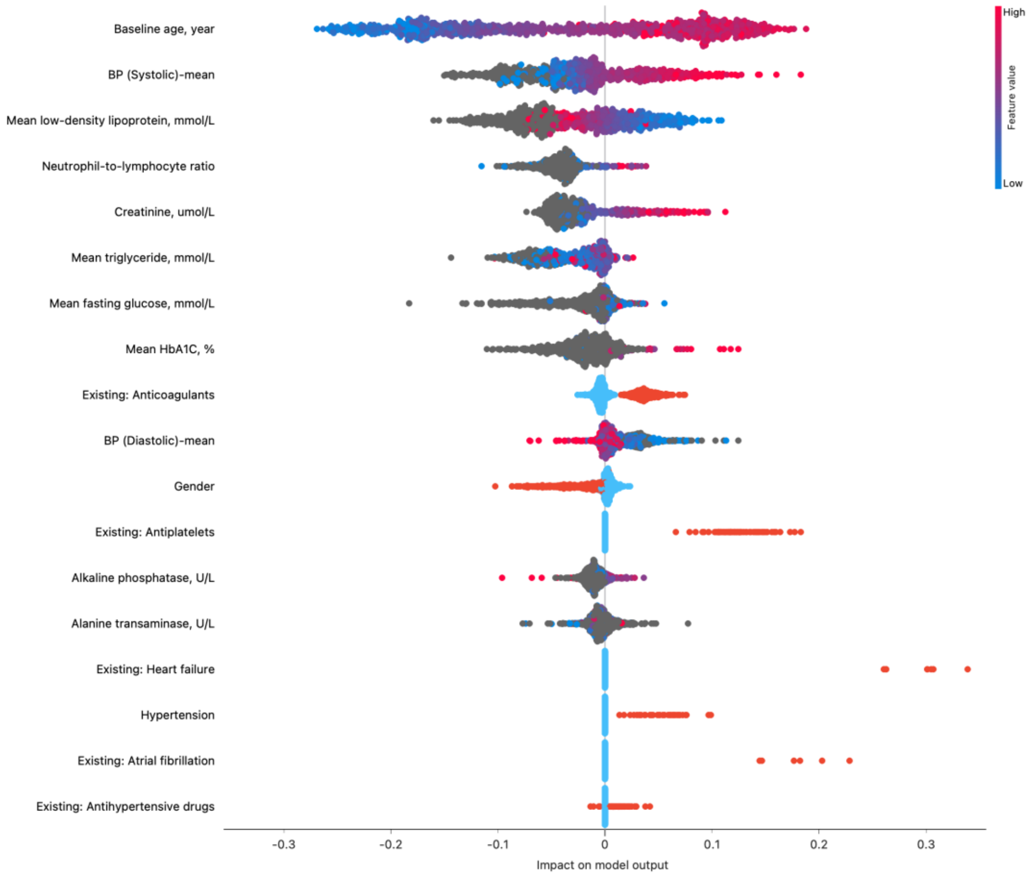

**Supplementary Figure 4.** SHapley Additive exPlanations (SHAP) feature importance interpretations for CatBoost model to predict MACE in patients attending family medicine clinics.

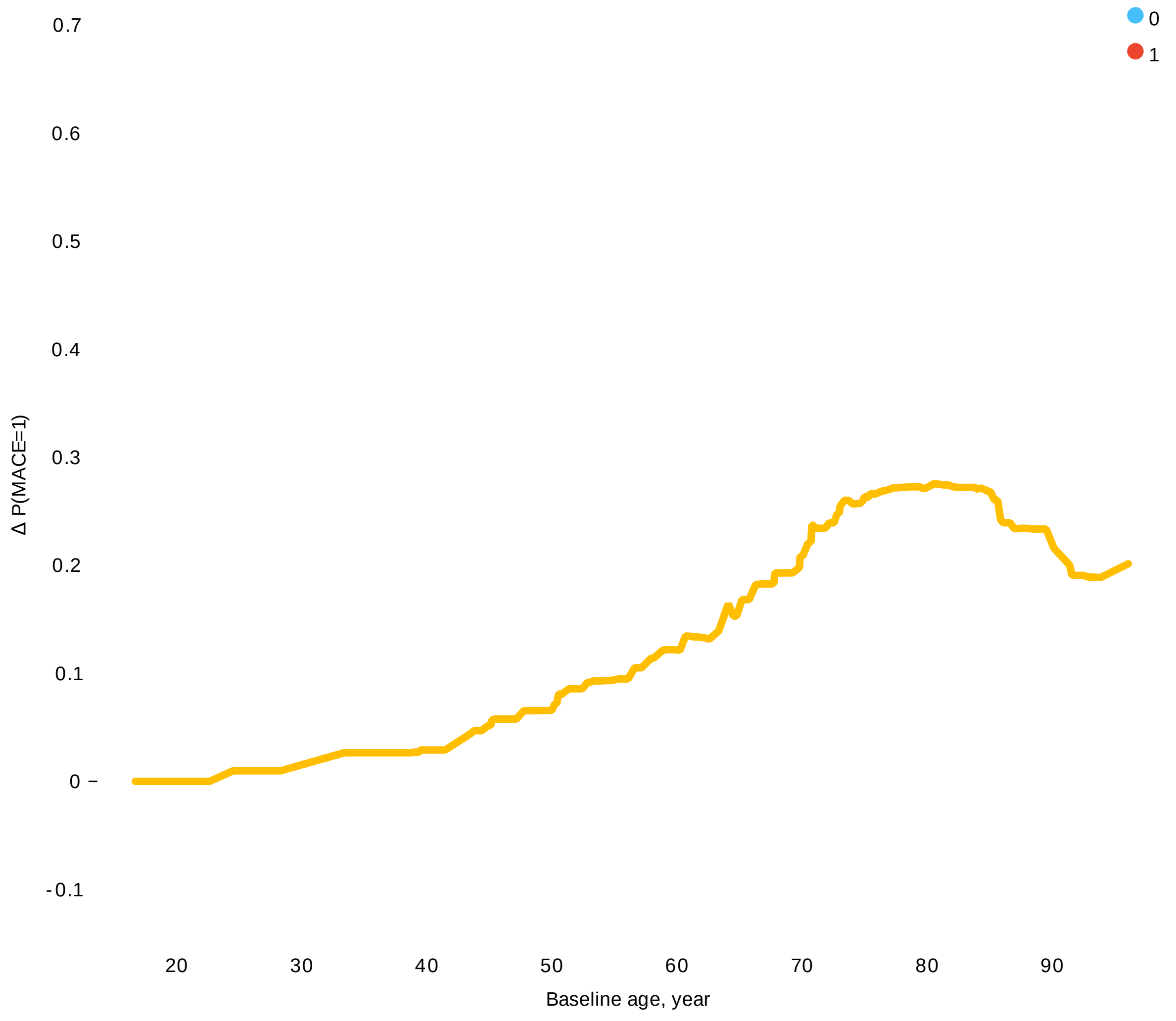

**Supplementary Figure 5.** Individual Conditional Expectation of age at admission to predict MACE in patients attending family medicine clinics.

Each line per patient is used to show how the prediction changes when age at admission changes).

**Supplementary Table 1.** The International Classification of Diseases, Clinical Modification (ICD-9) and (ICD-10) codes for definitions of past comorbidities and outcomes.

| |
|---|
| **Adverse outcome of interest** |
| MACE:<br>**1.** cardiovascular mortality<br>**2.** myocardial infarction<br>**3.** heart failure<br>**4.** stroke/ transient ischaemic attack |
| **Cardiovascular mortality:** ICD-10: I00-I78 |
| **Acute myocardial infarction:** 410 410.01 410.02 410.1 410.11 410.12 410.2 410.21 410.22 410.3 410.31 410.32 410.4 410.41 410.42 410.5 410.51 410.52 410.6 410.61 410.62 410.7 410.71 410.72 410.8 410.81 410.82 410.9 410.91 410.92 |
| **Heart failure:** 428 428 428.1 428.2 428.2 428.21 428.22 428.23 428.3 428.3 428.31 428.32 428.33 428.4 428.4 428.41 428.42 428.43 428.9 398.91 402.01 402.11 402.91 404.01 404.03 404.11 404.13 404.91 404.93 |
| **Stroke/transient ischemic attack:** 435 435.1 435.2 435.3 435.8 435.9 433.81 433.91 434 436 437 437.1 433.31 433.01 434.01 434.1 434.11 434.9 434.91 437.2 437.3 437.4 437.5 437.6 437.7 437.8 437.9 430 431 432 432.1 432.9 |
| **Past comorbidities** |
| **Hypertension**: 401 401.1 401.9 402 402.01 402.1 402.11 402.9 402.91 403 403.01 403.1 403.11 403.9 403.91 404 404.01 404.02 404.03 404.1 404.11 404.12 404.13 404.9 404.91 404.92 404.93 405 405.01 405.09 405.1 405.11 405.19 405.9 405.91 405.99 437.2 + history of uses of anti-hypertensives |
| Diabetes mellitus 250 250.01 250.02 250.03 250.1 250.11 250.12 250.13 250.2 250.21 250.22 250.23 250.3 250.31 250.32 250.33 250.4 250.41 250.42 250.43 250.5 250.51 250.52 250.53 250.6 250.61 250.62 250.63 250.7 250.71 250.72 250.73 250.8 250.81 250.82 250.83 250.9 250.91 250.92 250.93 |
| **Atrial fibrillation:** 427.31 429.4 |
| **Ischemic heart disease:** 410.01 410.02 410.1 410.11 410.12 410.2 410.21 410.22 410.3 410.31 410.32 410.4 410.41 410.42 410.5 410.51 410.52 410.6 410.61 410.62 410.7 410.71 410.72 410.8 410.81 410.82 410.9 410.91 410.92 411 411.1 411.8 411.81 411.89 413 413.1 413.9 414 414.01 414.02 414.03 414.04 414.05 414.06 414.07 414.1 414.11 414.12 414.19 414.2 414.3 414.4 414.8 414.9 410 412 |
| **Chronic obstructive pulmonary disease** 491.0 491.1 491.2 491.8 491.9 492.0 492.8 496 |

**Supplementary Table 2.** Univariable Cox regression to predict specific MACE events and all-cause mortality in patients attending family medicine clinics.
MACE: Major adverse cardiovascular events; TIA: transient ischemic attack; BP: Blood pressure; HR: Hazard ratio; CI: Confidence interval.

| Characteristics | All-cause mortality HR [95% CI];P value | Cardiovascular mortality HR [95% CI];P value | New onset myocardial infarction HR [95% CI];P value | New onset heart failure HR [95% CI];P value | New onset stroke/TIA HR [95% CI];P value |
|---|---|---|---|---|---|
| ***Demographics*** | | | | | |
| Male gender | 1.38[1.36-1.40];<0.0001*** | 1.18[1.14-1.23];<0.0001*** | 1.24[1.18-1.30];<0.0001*** | 0.93[0.90-0.97];0.0001*** | 1.13[1.08-1.17];<0.0001*** |
| Female gender | 0.72[0.71-0.74];<0.0001*** | 0.84[0.81-0.88];<0.0001*** | 0.81[0.77-0.85];<0.0001*** | 1.07[1.04-1.11];0.0001*** | 0.89[0.85-0.92];<0.0001*** |
| Baseline age, year | 1.056[1.056-1.057];<0.0001*** | 1.059[1.057-1.060];<0.0001*** | 1.041[1.039-1.043];<0.0001*** | 1.057[1.055-1.058];<0.0001*** | 1.030[1.029-1.032];<0.0001*** |
| 18-40 | 0.06[0.05-0.07];<0.0001*** | 0.07[0.05-0.08];<0.0001*** | 0.11[0.09-0.14];<0.0001*** | 0.05[0.04-0.06];<0.0001*** | 0.09[0.07-0.11];<0.0001*** |
| 40-50 | 0.21[0.20-0.22];<0.0001*** | 0.18[0.16-0.20];<0.0001*** | 0.25[0.22-0.27];<0.0001*** | 0.11[0.10-0.12];<0.0001*** | 0.33[0.30-0.35];<0.0001*** |
| 50-60 | 0.39[0.38-0.40];<0.0001*** | 0.34[0.32-0.36];<0.0001*** | 1.0[Reference] | 1.0[Reference] | 1.0[Reference] |
| 60-70 | 1.0[Reference] | 1.0[Reference] | 1.30[1.23-1.37];<0.0001*** | 1.06[1.01-1.10];0.0077** | 1.52[1.46-1.59];<0.0001*** |
| 70-80 | 2.46[2.42-2.50];<0.0001*** | 2.55[2.45-2.64];<0.0001*** | 3.03[2.88-3.18];<0.0001*** | 3.36[3.25-3.47];<0.0001*** | 2.41[2.32-2.51];<0.0001*** |
| >80 | 2.95[2.90-3.00];<0.0001*** | 3.04[2.92-3.17];<0.0001*** | 1.34[1.25-1.43];<0.0001*** | 2.17[2.09-2.26];<0.0001*** | 0.90[0.84-0.96];0.0013** |
| ***Comorbidities*** | | | | | |
| Diabetes mellitus | 1.34[1.30-1.38];<0.0001*** | 1.60[1.51-1.70];<0.0001*** | 3.00[2.82-3.19];<0.0001*** | 2.59[2.47-2.71];<0.0001*** | 2.08[1.97-2.20];<0.0001*** |
| Hypertension | 1.92[1.89-1.95];<0.0001*** | 2.11[2.04-2.19];<0.0001*** | 3.28[3.12-3.44];<0.0001*** | 3.84[3.71-3.97];<0.0001*** | 2.76[2.66-2.87];<0.0001*** |

| | | | | | |
|---|---|---|---|---|---|
| Chronic obstructive pulmonary disease | 6.27[5.80-6.76];<0.0001*** | 2.78[2.15-3.58];<0.0001*** | 4.76[3.63-6.24];<0.0001*** | 7.35[6.35-8.51];<0.0001*** | 2.10[1.50-2.92];<0.0001*** |
| Ischemic heart disease | 2.27[2.15-2.39];<0.0001*** | 3.15[2.85-3.49];<0.0001*** | 4.67[4.17-5.23];<0.0001*** | 6.01[5.59-6.45];<0.0001*** | 2.86[2.55-3.21];<0.0001*** |
| Heart failure | 5.78[5.42-6.16];<0.0001*** | 8.40[7.46-9.46];<0.0001*** | 7.21[6.03-8.62];<0.0001*** | 30.64[28.45-33.00];<0.0001*** | 3.78[3.10-4.61];<0.0001*** |
| Myocardial infarction | 2.49[2.14-2.89];<0.0001*** | 3.24[2.40-4.39];<0.0001*** | 6.55[4.88-8.78];<0.0001*** | 6.74[5.51-8.24];<0.0001*** | 2.45[1.67-3.60];<0.0001*** |
| Atrial fibrillation | 4.41[4.12-4.73];<0.0001*** | 6.77[5.95-7.69];<0.0001*** | 4.49[3.64-5.55];<0.0001*** | 14.20[12.97-15.56];<0.0001*** | 5.74[4.90-6.73];<0.0001*** |
| Stroke/TIA | 2.39[2.20-2.60];<0.0001*** | 3.09[2.60-3.67];<0.0001*** | 3.68[2.98-4.55];<0.0001*** | 3.46[2.98-4.02];<0.0001*** | 8.07[7.13-9.13];<0.0001*** |
| ***Medications*** | | | | | |
| Anticoagulants | 1.57[1.55-1.60];<0.0001*** | 2.03[1.95-2.10];<0.0001*** | 2.43[2.31-2.55];<0.0001*** | 2.71[2.62-2.81];<0.0001*** | 1.91[1.84-1.99];<0.0001*** |
| Antiplatelets | 2.28[2.23-2.33];<0.0001*** | 3.27[3.12-3.42];<0.0001*** | 3.49[3.29-3.70];<0.0001*** | 3.70[3.55-3.85];<0.0001*** | 3.16[3.01-3.31];<0.0001*** |
| Antihypertensive drugs | 2.04[2.01-2.08];<0.0001*** | 2.19[2.11-2.28];<0.0001*** | 2.59[2.46-2.72];<0.0001*** | 2.98[2.88-3.09];<0.0001*** | 2.12[2.03-2.21];<0.0001*** |
| Statins | 1.09[1.06-1.12];<0.0001*** | 1.46[1.38-1.55];<0.0001*** | 2.31[2.16-2.47];<0.0001*** | 1.93[1.83-2.03];<0.0001*** | 1.83[1.73-1.93];<0.0001*** |
| ***Laboratory tests*** | | | | | |
| Neutrophil-to-lymphocyte ratio | 1.03[1.02-1.03];<0.0001*** | 1.02[1.02-1.03];<0.0001*** | 1.02[1.02-1.03];<0.0001*** | 1.02[1.02-1.03];<0.0001*** | 1.02[1.01-1.02];<0.0001*** |
| Creatinine, umol/L | 1.003[1.003-1.003];<0.0001*** | 1.003[1.003-1.004];<0.0001*** | 1.003[1.003-1.004];<0.0001*** | 1.003[1.003-1.003];<0.0001*** | 1.003[1.003-1.003];<0.0001*** |
| Alkaline phosphatase, U/L | 1.001[1.001-1.001];<0.0001*** | 1.001[1.001-1.001];<0.0001*** | 1.001[1.001-1.001];<0.0001*** | 1.001[1.001-1.001];<0.0001*** | 1.001[1.000-1.001];<0.0001*** |

| | | | | | |
|---|---|---|---|---|---|
| Aspartate transaminase, U/L | 1.000[1.000-1.000];0.4337 | 1.000[1.000-1.001];0.7063 | 1.000[1.000-1.001];0.0880 | 1.000[0.999-1.000];0.2494 | 0.999[0.998-1.000];0.1421 |
| Alanine transaminase, U/L | 0.999[0.998-0.999];<0.0001*** | 1.00[0.99-1.00];<0.0001*** | 0.998[0.997-1.000];0.0296* | 0.998[0.997-0.999];0.0007*** | 0.999[0.998-1.000];0.1734 |
| BP (Systolic)-mean | 1.029[1.029-1.030];<0.0001*** | 1.03[1.03-1.04];<0.0001*** | 1.030[1.028-1.032];<0.0001*** | 1.032[1.031-1.033];<0.0001*** | 1.02[1.02-1.03];<0.0001*** |
| BP (Diastolic)-mean | 0.98[0.97-0.98];<0.0001*** | 0.978[0.976-0.981];<0.0001*** | 0.98[0.97-0.98];<0.0001*** | 0.97[0.96-0.97];<0.0001*** | 0.99[0.99-1.00];<0.0001*** |
| Mean HbA1C, % | 1.05[1.03-1.06];<0.0001*** | 1.09[1.05-1.13];<0.0001*** | 1.13[1.09-1.17];<0.0001*** | 1.09[1.06-1.12];<0.0001*** | 1.06[1.03-1.09];0.0004*** |
| Mean fasting glucose, mmol/L | 1.08[1.08-1.09];<0.0001*** | 1.09[1.08-1.10];<0.0001*** | 1.10[1.09-1.12];<0.0001*** | 1.08[1.08-1.09];<0.0001*** | 1.06[1.05-1.07];<0.0001*** |
| Mean triglyceride, mmol/L | 0.99[0.97-1.00];0.0255* | 1.04[1.02-1.07];0.0009*** | 1.03[1.00-1.06];0.0608 | 0.91[0.89-0.94];<0.0001*** | 0.93[0.91-0.96];<0.0001*** |
| Mean low-density lipoprotein, mmol/L | 1.01[1.00-1.03];0.1050 | 1.03[1.00-1.07];0.0556 | 0.62[0.60-0.64];<0.0001*** | 0.66[0.64-0.68];<0.0001*** | 0.62[0.60-0.64];<0.0001*** |
| Mean high-density lipoprotein, mmol/L | 0.60[0.58-0.62];<0.0001*** | 0.49[0.45-0.53];<0.0001*** | 0.33[0.30-0.36];<0.0001*** | 0.56[0.53-0.59];<0.0001*** | 0.56[0.53-0.59];<0.0001*** |

**Supplementary Table 3.** Univariable Cox regression to predict MACE in patients attending family medicine clinics.
MACE: Major adverse cardiovascular events; TIA: transient ischemic attack; BP: Blood pressure; HR: Hazard ratio; CI: Confidence interval.

| Characteristics | MACE HR [95% CI];P value | Optimal Cutoff |
|---|---|---|
| ***Demographics*** | | |
| Male gender | 1.13[1.11-1.16];<0.0001*** | - |
| Female gender | 0.88[0.87-0.90];<0.0001*** | - |
| Baseline age, year | 1.04[1.04-1.05];<0.0001*** | 62.18 |
| 18-40 | 0.08[0.07-0.09];<0.0001*** | - |
| 40-50 | 0.23[0.22-0.25];<0.0001*** | - |
| 50-60 | 0.47[0.45-0.48];<0.0001*** | - |
| 60-70 | 1.0[Reference] | - |
| 70-80 | 2.68[2.62-2.75];<0.0001*** | - |
| >80 | 1.86[1.81-1.91];<0.0001*** | - |
| ***Comorbidities*** | | |
| Diabetes mellitus | 2.07[2.00-2.14];<0.0001*** | - |
| Hypertension | 2.76[2.70-2.82];<0.0001*** | - |
| Chronic obstructive pulmonary disease | 4.67[4.12-5.30];<0.0001*** | - |
| Ischemic heart disease | 4.04[3.81-4.29];<0.0001*** | - |
| Heart failure | 15.44[14.42-16.54];<0.0001*** | - |
| Myocardial infarction | 4.39[3.70-5.21];<0.0001*** | - |
| Atrial fibrillation | 9.25[8.56-10.00];<0.0001*** | - |
| Stroke/TIA | 4.64[4.23-5.09];<0.0001*** | - |
| ***Medications*** | | |
| Anticoagulants | 2.15[2.11-2.20];<0.0001*** | - |
| Antiplatelets | 3.33[3.24-3.42];<0.0001*** | - |
| Antihypertensive drugs | 2.38[2.33-2.44];<0.0001*** | - |
| Statins | 1.75[1.69-1.81];<0.0001*** | - |
| ***Laboratory tests*** | | |
| Neutrophil-to-lymphocyte ratio | 1.022[1.020-1.025];<0.0001*** | 2.86 |
| Creatinine, umol/L | 1.003[1.003-1.003];<0.0001*** | 104 |
| Alkaline phosphatase, U/L | 1.001[1.001-1.001];<0.0001*** | 72 |
| Aspartate transaminase, U/L | 1.000[0.999-1.000];0.4357 | 12.386 |
| Alanine transaminase, U/L | 0.999[0.998-0.999];0.0001*** | 19.858 |
| BP (Systolic)-mean | 1.029[1.029-1.030];<0.0001*** | 138.1864 |
| BP (Diastolic)-mean | 0.979[0.978-0.981];<0.0001*** | 72.3 |
| Mean HbA1C, % | 1.08[1.06-1.10];<0.0001*** | 7.9857 |
| Mean fasting glucose, mmol/L | 1.08[1.07-1.08];<0.0001*** | 5.6113 |
| Mean triglyceride, mmol/L | 0.97[0.95-0.98];<0.0001*** | 1.8494 |
| Mean low-density lipoprotein, mmol/L | 0.69[0.67-0.70];<0.0001*** | 2.5412 |
| Mean high-density lipoprotein, mmol/L | 0.53[0.51-0.55];<0.0001*** | 1.1338 |

**Supplementary Table 4.** Distribution analysis for the CatBoost model predicted probability as an *in-silico* marker to predict MACE in patients attending family medicine clinics.
MACE: Major adverse cardiovascular events; IQR: Interquartile range.

| **MACE (N=31601)**<br>**Median (IQR);N** | **No MACE (N=122968)**<br>**Median (IQR);N** | **P value** |
|---|---|---|
| 0.41(0.19-0.66);n=31601 | 0.08(0.03-0.19);n=122968 | <0.0001*** |
| **Cardiovascular mortality (N=11190)**<br>**Median (IQR);N** | **No cardiovascular mortality (N=143379)**<br>**Median (IQR);N** | **P value** |
| 0.31(0.12-0.61);n=11190 | 0.1(0.04-0.27);n=143379 | <0.0001*** |
| **New onset myocardial infarction (N=6704)**<br>**Median (IQR);N** | **No new onset myocardial infarction (N=147865)**<br>**Median (IQR);N** | **P value** |
| 0.49(0.27-0.7);n=6704 | 0.1(0.04-0.26);n=147865 | <0.0001*** |
| **New onset heart failure (N=13826)**<br>**Median (IQR);N** | **No new onset heart failure (N=140743)**<br>**Median (IQR);N** | **P value** |
| 0.52(0.3-0.73);n=13826 | 0.09(0.04-0.23);n=140743 | <0.0001*** |
| **New onset stroke/TIA (N=10446)**<br>**Median (IQR);N** | **No New onset stroke/TIA (N=144123)**<br>**Median (IQR);N** | **P value** |
| 0.43(0.23-0.66);n=10446 | 0.09(0.04-0.25);n=144123 | <0.0001*** |

**Supplementary Table 5.** Prediction strength of the CatBoost model predicted probability as an *in-silico* marker to predict MACE in patients attending family medicine clinics with different follow-up durations from the baseline recruitment date.
MACE: Major adverse cardiovascular events; HR: Hazard ratio; CI: Confidence interval.

| **Follow-up duration, years** | **MACE HR [95% CI];P value** | **Cardiovascular mortality HR [95% CI];P value** | **New onset myocardial infarction HR [95% CI];P value** | **New onset heart failure HR [95% CI];P value** | **New onset stroke/TIA HR [95% CI];P value** |
|---|---|---|---|---|---|
| <1 | 5.18[4.45-6.02];<0.0001*** | 0.94[0.74-1.18];0.5853 | 2.15[1.52-3.05];<0.0001*** | 9.58[7.80-11.76];<0.0001*** | 1.80[1.21-2.66];0.0034** |
| 1-2 | 5.09[4.34-5.97];<0.0001*** | 0.85[0.66-1.11];0.2345 | 3.82[2.66-5.50];<0.0001*** | 7.42[5.93-9.28];<0.0001*** | 2.58[1.69-3.94];<0.0001*** |
| 2-3 | 5.53[4.72-6.49];<0.0001*** | 0.86[0.66-1.11];0.2447 | 3.16[2.18-4.57];<0.0001*** | 12.33[9.75-15.59];<0.0001*** | 2.44[1.75-3.39];<0.0001*** |
| 3-4 | 4.91[4.23-5.70];<0.0001*** | 0.89[0.69-1.14];0.3377 | 5.64[3.84-8.29];<0.0001*** | 9.08[7.16-11.52];<0.0001*** | 3.12[2.38-4.09];<0.0001*** |
| 4-5 | 4.68[4.04-5.43];<0.0001*** | 0.66[0.50-0.85];0.0017** | 4.31[3.00-6.19];<0.0001*** | 8.65[6.81-10.98];<0.0001*** | 3.25[2.54-4.15];<0.0001*** |
| 5-6 | 5.05[4.37-5.84];<0.0001*** | 0.73[0.56-0.94];0.0144* | 5.23[3.68-7.44];<0.0001*** | 6.01[4.78-7.55];<0.0001*** | 4.33[3.40-5.52];<0.0001*** |
| 6-7 | 6.41[5.50-7.47];<0.0001*** | 0.64[0.49-0.84];0.0013** | 3.81[2.65-5.49];<0.0001*** | 8.49[6.70-10.78];<0.0001*** | 4.33[3.32-5.66];<0.0001*** |
| 7-8 | 4.80[4.13-5.59];<0.0001*** | 0.74[0.57-0.97];0.0271* | 4.30[2.97-6.21];<0.0001*** | 8.49[6.70-10.75];<0.0001*** | 3.81[2.90-4.99];<0.0001*** |
| 8-9 | 4.98[4.27-5.80];<0.0001*** | 0.68[0.51-0.90];0.0067** | 2.94[2.06-4.18];<0.0001*** | 8.68[6.88-10.94];<0.0001*** | 3.83[2.90-5.06];<0.0001*** |
| 9-10 | 5.53[4.70-6.49];<0.0001*** | 0.75[0.55-1.02];0.0658 | 3.38[2.38-4.79];<0.0001*** | 9.23[7.21-11.80];<0.0001*** | 3.61[2.67-4.87];<0.0001*** |
| 10-11 | 5.17[4.35-6.13];<0.0001*** | 0.73[0.52-1.02];0.0630 | 4.28[3.00-6.10];<0.0001*** | 9.91[7.63-12.86];<0.0001*** | 3.40[2.43-4.75];<0.0001*** |
| 11-12 | 5.99[5.01-7.16];<0.0001*** | 0.79[0.56-1.12];0.1929 | 5.04[3.53-7.21];<0.0001*** | 10.95[8.28-14.49];<0.0001*** | 3.86[2.78-5.38];<0.0001*** |

| | | | | | |
|---|---|---|---|---|---|
| 12-13 | 5.00[4.18-5.97];<0.0001*** | 0.84[0.59-1.20];0.3420 | 4.62[3.27-6.52];<0.0001*** | 8.44[6.44-11.05];<0.0001*** | 2.48[1.74-3.52];<0.0001*** |
| 13-14 | 5.11[4.21-6.20];<0.0001*** | 0.88[0.62-1.26];0.4960 | 6.35[4.32-9.31];<0.0001*** | 7.87[5.82-10.64];<0.0001*** | 2.98[2.02-4.40];<0.0001*** |
| 14-15 | 6.25[5.14-7.60];<0.0001*** | 0.57[0.39-0.85];0.0058** | 7.19[4.97-10.40];<0.0001*** | 8.41[6.17-11.46];<0.0001*** | 4.06[2.76-5.99];<0.0001*** |
| 15-16 | 52.28[44.24-61.78];<0.0001*** | 5.73[3.90-8.41];<0.0001*** | 43.39[29.90-62.97];<0.0001*** | 67.31[50.38-89.93];<0.0001*** | 13.54[10.29-17.83];<0.0001*** |
| 16-17 | 47.69[40.23-56.53];<0.0001*** | 14.92[10.05-22.16];<0.0001*** | 54.44[36.69-80.77];<0.0001*** | 59.48[43.79-80.80];<0.0001*** | 19.85[15.34-25.71];<0.0001*** |
| 17-18 | 48.49[40.18-58.52];<0.0001*** | 11.61[6.99-19.28];<0.0001*** | 44.87[26.12-77.09];<0.0001*** | 52.14[35.91-75.71];<0.0001*** | 34.89[27.21-44.74];<0.0001*** |
| 18-19 | 51.33[39.38-66.90];<0.0001*** | 8.10[3.69-17.80];<0.0001*** | 36.76[14.43-93.64];<0.0001*** | 50.98[28.42-91.46];<0.0001*** | 61.43[43.99-85.77];<0.0001*** |
| 19-20 | 44.69[25.71-77.68];<0.0001*** | 65.07[10.72-394.85];<0.0001*** | 16.84[3.30-85.88];0.0007*** | 22.34[5.92-84.31];<0.0001*** | 48.81[25.45-93.61];<0.0001*** |
